# Polygenic Risk Modelling for Prediction of Epithelial Ovarian Cancer Risk

**DOI:** 10.1101/2020.11.30.20219220

**Authors:** Eileen O. Dareng, Jonathan P. Tyrer, Daniel R. Barnes, Michelle R. Jones, Xin Yang, Katja K.H. Aben, Muriel A. Adank, Simona Agata, Irene L. Andrulis, Hoda Anton-Culver, Natalia N. Antonenkova, Gerasimos Aravantinos, Banu K. Arun, Annelie Augustinsson, Judith Balmaña, Elisa V. Bandera, Rosa B. Barkardottir, Daniel Barrowdale, Matthias W. Beckmann, Alicia Beeghly-Fadiel, Javier Benitez, Marina Bermisheva, Marcus Q. Bernardini, Line Bjorge, Amanda Black, Natalia V. Bogdanova, Bernardo Bonanni, Ake Borg, James D. Brenton, Agnieszka Budzilowska, Ralf Butzow, Saundra S. Buys, Hui Cai, Maria A. Caligo, Ian Campbell, Rikki Cannioto, Hayley Cassingham, Jenny Chang-Claude, Stephen J. Chanock, Kexin Chen, Yoke-Eng Chiew, Wendy K. Chung, Kathleen B.M. Claes, Sarah Colanna, GEMO Study Collaborators, GC-HBOC study Collaborators, EMBRACE Collaborators, Linda S. Cook, Fergus J. Couch, Mary B. Daly, Fanny Dao, Eleanor Davies, Miguel de la Hoya, Robin de Putter, Joe Dennis, Allison DePersia, Peter Devilee, Orland Diez, Yuan Chun Ding, Jennifer A. Doherty, Susan M. Domchek, Thilo Dörk, Andreas du Bois, Matthias Dürst, Diana M. Eccles, Heather A. Eliassen, Christoph Engel, D. Gareth Evans, Peter A. Fasching, James M. Flanagan, Lenka Foretova, Renée T. Fortner, Eitan Friedman, Patricia A. Ganz, Judy Garber, Francesca Gensini, Graham G. Giles, Gord Glendon, Andrew K. Godwin, Marc T. Goodman, Mark H. Greene, Jacek Gronwald, OPAL Study Group, AOCS Group, Eric Hahnen, Christopher A. Haiman, Niclas Håkansson, Ute Hamann, Thomas V.O. Hansen, Holly R. Harris, Mikael Hartman, Florian Heitz, Michelle A.T. Hildebrandt, Estrid Høgdall, Claus K. Høgdall, John L. Hopper, Ruea-Yea Huang, Chad Huff, Peter J. Hulick, David G. Huntsman, Evgeny N. Imyanitov, KConFab Investigators, HEBON Investigators, Claudine Isaacs, Anna Jakubowska, Paul A. James, Ramunas Janavicius, Allan Jensen, Oskar Th. Johannsson, Esther M. John, Michael E. Jones, Daehee Kang, Beth Y. Karlan, Anthony Karnezis, Linda E. Kelemen, Elza Khusnutdinova, Lambertus A. Kiemeney, Byoung-Gie Kim, Susanne K. Kjaer, Ian Komenaka, Jolanta Kupryjanczyk, Allison W. Kurian, Ava Kwong, Diether Lambrechts, Melissa C. Larson, Conxi Lazaro, Nhu D. Le, Goska Leslie, Jenny Lester, Fabienne Lesueur, Douglas A. Levine, Lian Li, Jingmei Li, Jennifer T. Loud, Karen H. Lu, Jan Lubiński, Eva Machackova, Phuong L. Mai, Siranoush Manoukian, Jeffrey R. Marks, Rayna Kim Matsuno, Keitaro Matsuo, Taymaa May, Lesley McGuffog, John R. McLaughlin, Iain A. McNeish, Noura Mebirouk, Usha Menon, Austin Miller, Roger L. Milne, Albina Minlikeeva, Francesmary Modugno, Marco Montagna, Kirsten B. Moysich, Elizabeth Munro, Katherine L. Nathanson, Susan L. Neuhausen, Heli Nevanlinna, Joanne Ngeow Yuen Yie, Henriette Roed Nielsen, Finn C. Nielsen, Liene Nikitina-Zake, Kunle Odunsi, Kenneth Offit, Edith Olah, Siel Olbrecht, Olufunmilayo I. Olopade, Sara H. Olson, Håkan Olsson, Ana Osorio, Laura Papi, Sue K. Park, Michael T. Parsons, Harsha Pathak, Inge Sokilde Pedersen, Ana Peixoto, Tanja Pejovic, Pedro Perez-Segura, Jennifer B. Permuth, Beth Peshkin, Paolo Peterlongo, Anna Piskorz, Darya Prokofyeva, Paolo Radice, Johanna Rantala, Marjorie J. Riggan, Harvey A. Risch, Cristina Rodriguez-Antona, Eric Ross, Mary Anne Rossing, Ingo Runnebaum, Dale P. Sandler, Marta Santamariña, Penny Soucy, Rita K. Schmutzler, V. Wendy Setiawan, Kang Shan, Weiva Sieh, Jacques Simard, Christian F. Singer, Anna P Sokolenko, Honglin Song, Melissa C. Southey, Helen Steed, Dominique Stoppa-Lyonnet, Rebecca Sutphen, Anthony J. Swerdlow, Yen Yen Tan, Manuel R. Teixeira, Soo Hwang Teo, Kathryn L. Terry, Mary Beth Terry, Mads Thomassen, Pamela J. Thompson, Liv Cecilie Vestrheim Thomsen, Darcy L. Thull, Marc Tischkowitz, Linda Titus, Amanda E. Toland, Diana Torres, Britton Trabert, Ruth Travis, Nadine Tung, Shelley S. Tworoger, Ellen Valen, Anne M. van Altena, Annemieke H. van der Hout, Els Van Nieuwenhuysen, Elizabeth J. van Rensburg, Ana Vega, Digna Velez Edwards, Robert A. Vierkant, Frances Wang, Barbara Wappenschmidt, Penelope M. Webb, Clarice R. Weinberg, Jeffrey N. Weitzel, Nicolas Wentzensen, Emily White, Alice S. Whittemore, Stacey J. Winham, Alicja Wolk, Yin-Ling Woo, Anna H. Wu, Li Yan, Drakoulis Yannoukakos, Katia M. Zavaglia, Wei Zheng, Argyrios Ziogas, Kristin K. Zorn, Douglas Easton, Kate Lawrenson, Anna DeFazio, Thomas A. Sellers, Susan J. Ramus, Celeste L. Pearce, Alvaro N. Monteiro, Julie Cunningham, Ellen L. Goode, Joellen M. Schildkraut, Andrew Berchuck, Georgia Chenevix-Trench, Simon A. Gayther, Antonis C. Antoniou, Paul D.P. Pharoah

## Abstract

Polygenic risk scores (PRS) for epithelial ovarian cancer (EOC) have the potential to improve risk stratification. Joint estimation of Single Nucleotide Polymorphism (SNP) effects in models could improve predictive performance over standard approaches of PRS construction. Here, we implemented computationally-efficient, penalized, logistic regression models (lasso, elastic net, stepwise) to individual level genotype data and a Bayesian framework with continuous shrinkage, “select and shrink for summary statistics” (S4), to summary level data for epithelial non-mucinous ovarian cancer risk prediction. We developed the models in a dataset consisting of 23,564 non-mucinous EOC cases and 40,138 controls participating in the Ovarian Cancer Association Consortium (OCAC) and validated the best models in three populations of different ancestries: prospective data from 198,101 women of European ancestry; 7,669 women of East Asian ancestry; 1,072 women of African ancestry, and in 18,915 *BRCA1* and 12,337 *BRCA2* pathogenic variant carriers of European ancestry. In the external validation data, the model with the strongest association for non-mucinous EOC risk derived from the OCAC model development data was the S4 model (27,240 SNPs) with odds ratios (OR) of 1.38(95%CI:1.28–1.48,AUC:0.588) per unit standard deviation, in women of European ancestry; 1.14(95%CI:1.08–1.19,AUC:0.538) in women of East Asian ancestry; 1.38(95%CI:1.21-1.58,AUC:0.593) in women of African ancestry; hazard ratios of 1.37(95%CI:1.30–1.44,AUC:0.592) in *BRCA1* pathogenic variant carriers and 1.51(95%CI:1.36-1.67,AUC:0.624) in *BRCA2* pathogenic variant carriers. Incorporation of the S4 PRS in risk prediction models for ovarian cancer may have clinical utility in ovarian cancer prevention programs.

## INTRODUCTION

Rare mutations in known high and moderate penetrance susceptibility genes (*BRCA1, BRCA2, BRIP1, PALB2, RAD51C, RAD51D* and the mis-match repair genes) account for about 40 percent of the inherited component of EOC disease risk (1,2). Genome wide association studies (GWAS), reviewed in Kar et. al. and Jones et. al. (1,3), have identified 39 common (minor allele frequency [MAF] > 0.05) susceptibility variants which together explain about 6% of the heritability of EOC. Polygenic risk scores (PRS) provide an opportunity for refined risk stratification in the general population as well as in carriers of rare moderate or high risk alleles (4,5).

A PRS is calculated as the weighted sum of the number of risk alleles carried for a specified set of genetic variants. The best approach to identify the set of variants and their weights in order to optimize the predictive power of a PRS is unknown. A common approach involves selecting a set of variants that reach a threshold for association based on the p-value for each variant with or without clumping and pruning to remove highly correlated variants (6,7). More complex prediction models, based on machine learning approaches that do not assume variant independence have also been used to construct PRS for complex traits in humans (8,9). To date, these methods have produced only modest gains in predictive power for highly polygenic phenotypes (8,10). Penalized regression approaches such as the lasso, elastic net and the adaptive lasso have also been used for the joint estimation of variant effects using individual level data for large data sets (11). While they have the potential advantage of improving performance, the major drawback of these methods is the high computational burden required to fit the models (11,12).

In this study, we present a novel implementation of computationally-efficient PRS models using two approaches: 1) penalized regression models including the lasso, elastic net and minimax concave penalty, for use when individual genotype data are available; and 2) a Bayesian regression model with continuous shrinkage priors on variant effect sizes, for use in broader settings where summary statistics are available, hereafter referred to as the “select and shrink with summary statistics” (S4) method. We compare these models with two commonly used methods, stepwise regression with p-value thresholding and LDPred.

## MATERIALS (SUBJECTS) AND METHODS

### EOC Histotypes

EOC is a highly heterogeneous phenotype with five major histotypes for invasive disease – high-grade serous, low-grade serous, endometrioid, clear cell and mucinous histotype. The mucinous histotype is the least common and its origin is the most controversial with up to 60% of diagnosed cases of mucinous ovarian cancer often being misdiagnosed metastasis from non-ovarian sites (13). Recent molecular analyses have concluded that most primary invasive mucinous cases are not extra-ovarian metastases (14). However, accurate diagnosis relies on expert histopathology and immuno-histochemical profiling (15), which remains a challenge in clinical practice and can be an issue in different cohorts from different time periods. Therefore, in this study, we performed PRS modelling and association testing for all cases of invasive EOC, excluding mucinous cases, hereafter referred to as non-mucinous EOC.

### Model Development Study Population

We used genotype data from 23,564 invasive non-mucinous EOC cases and 40,138 controls with >80% European ancestry from 63 case-control studies included in the Ovarian Cancer Association Consortium (OCAC) for model development. The study protocol was approved by the institutional review boards of the Brigham and Women’s Hospital and Harvard T.H. Chan School of Public Health, and those of participating registries as required. The distribution of cases by histotype was high-grade serous (13,609), low-grade serous (2,749), endometrioid (2,877), clear cell (1,427), and others (2,902). All mucinous EOC histotypes (2,587) were excluded. Sample collection, genotyping and quality control have been previously described (16). Genotype data were imputed to the Haplotype Reference Consortium reference panel on the Michigan Imputation server, using 470,825 SNPs that passed quality control. Of the 32 million SNPs imputed, 10 million had imputation r^2^ > 0.3 and were included in this analysis.

### Model Validation Study Population

#### UK Biobank Population

We validated the best-fitting PRS models developed in the OCAC data in 657 prevalent and incident cases of invasive EOC (346 serous, 98 endometrioid, 51 clear cell and 162 other) and 198,101 female controls of European ancestry from the UK Biobank. As with the model development data, all mucinous histotypes (166) were excluded. Samples were genotyped using either the Affymetrix UK BiLEVE Axiom Array or Affymetrix UK Biobank Axiom Array (which share 95% marker content), and then imputed to a combination of the Haplotype Reference Consortium, the 1000Genomes phase 3 and the UK10K reference panels (17). We restricted analysis to genetically confirmed females of European/white British ancestry. We excluded individuals if they were outliers for heterozygosity, had low genotyping call rate <95%, had sex chromosome aneuploidy, or if they were duplicates (cryptic or intended) (16). All SNPs selected in the model development phase were available in the UK Biobank.

#### Non-European Ancestry Population

We investigated transferability of the best-fitting PRS models to populations of non-European ancestry using genotype data from females of East Asian and African ancestries that had been genotyped as part of the OCAC OncoArray Project (18,19). Women of East Asian ancestry - 2,841 non-mucinous invasive EOC (1,960 high-grade serous, 136 low-grade serous, 400 endometrioid, 271 clear cell, 74 other histotypes) and 4,828 controls - were identified using a criterion of >80% Asian ancestry. This group included samples collected from population-based studies in China, Japan, Korea, Malaysia as well as samples from studies conducted in the US, Europe and Australia. Details of these data have been previously described (18). Similarly, women of African ancestry - 368 cases of non-mucinous invasive EOC (261 high-grade serous, 35 low-grade serous, 47 endometrioid, 7 clear cell, 53 other histoptypes) and 704 controls, mainly from studies conducted in the US, were identified using a criterion of >80% African ancestry.as described previously (19).

#### *BRCA1/BRCA2* Pathogenic Variant Carrier Population

We also assessed the performance of the best-fitting PRS models in women of European ancestry (>80% European ancestry) with the pathogenic *BRCA1* and *BRCA2* variants from the Consortium of Investigators of Modifiers of *BRCA1/2* (CIMBA). We used genotype data from 18,915 *BRCA1* (2,053 invasive EOC cases – 712 serous, 115 endometrioid, 9 clear cell, 1217 unknown/other) and 12,337 *BRCA2* (717 invasive EOC cases – 26 serous, 4 endometrioid, 1 clear cell, 686 unknown/other) pathogenic variant carriers from 63 studies contributing to CIMBA for independent model validation. Details of the study population and sample collection have been described previously (16). Genotyping, data quality control measures, intercontinental ancestry assessment and imputation to the HRC reference panel are as described for the OCAC study population.

#### PRS from Meta-analysis of Summary Statistics

We leveraged the increase in sample size resulting from a meta-analysis of EOC risk associations, using both the CIMBA and OCAC data described above, to explore performance of PRS approaches based on summary statistics.

### Statistical Analysis

#### Polygenic Risk Models

For all PRS models, we created scores as linear functions of the allele dosage in the general form 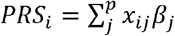 where genotypes are denoted as x (taking on the minor allele dosages of 0, 1 and 2), with *x*_*ij*_ representing the ith individual for the *j* th SNP (out of p SNPs) on an additive log scale and *β*_j_ represents the weight - the log of the odds ratio - of the *j* th SNP. We used different approaches to select and derive the optimal weights, *β*_*j*_, in models as described below.

#### Penalized logistic regression models: the lasso, elastic net and minimax concave penalty

A penalized logistic regression model for a set of SNPs aims to identify a set of regression coefficients that minimize the regularized loss function given by

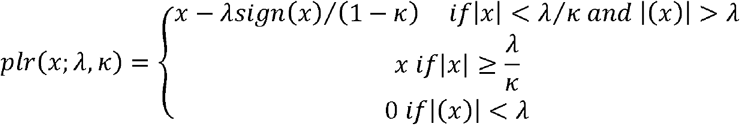

where *x* is the effect estimate of a SNP, *λ* is the tuning parameter and *κ* is the threshold (penalty) for different regularization paths. *λ* and *κ* are parameters that need to be chosen during model development to optimize performance. The lasso, elastic net, minimax concave penalty (MCP), and p-value thresholds are instances of the function with different *κ* values. We minimized the winner’s curse effect on inflated effect estimates for rare SNPs by penalizing rarer SNPs more heavily than common SNPs. Details are provided in the Supplementary Methods.

We used a two-stage approach to reduce computational burden without a corresponding loss in predictive power. The first stage was a SNP selection stage using a sliding windows approach, with 5.5Mb data blocks and a 500kb overlap between blocks. SNP selection was performed for each block and selected SNPs were collated. Single SNP association analyses were then run, and all SNPs with a χ^2^ test statistic of less than 2.25 were excluded. Penalized regression models were applied to the remaining SNPs using *λ* values of 3.0 and *κ* values of 0.0, 0.2, 0.4, 0.6, 0.8 and 1.0. SNPs selected in any of these models were included in subsequent analyses.

In the second stage, we fit penalized regression models to the training dataset with *λ* values ranging from 3.0 to 5.5 in increments of 0.1 iterated over *κ* values from −3.0 to 1 in increments of 0.1. The lasso model (*κ* = 0) for each value of *λ* was fitted first, to obtain a unique maximum. From the fitted maximum the *κ* value was changed, and the model refitted.

We applied this two-stage approach with five-fold cross-validation (**Figure 1**). The variants and their weights from the two-stage penalized logistic regression modelling in the training data were used to calculate the area under the receiver operating characteristic curve (AUC) in the test data. We repeated this process for each cross-validation iteration to obtain a mean AUC for each combination of *λ* and *κ*.

**Figure 1:**
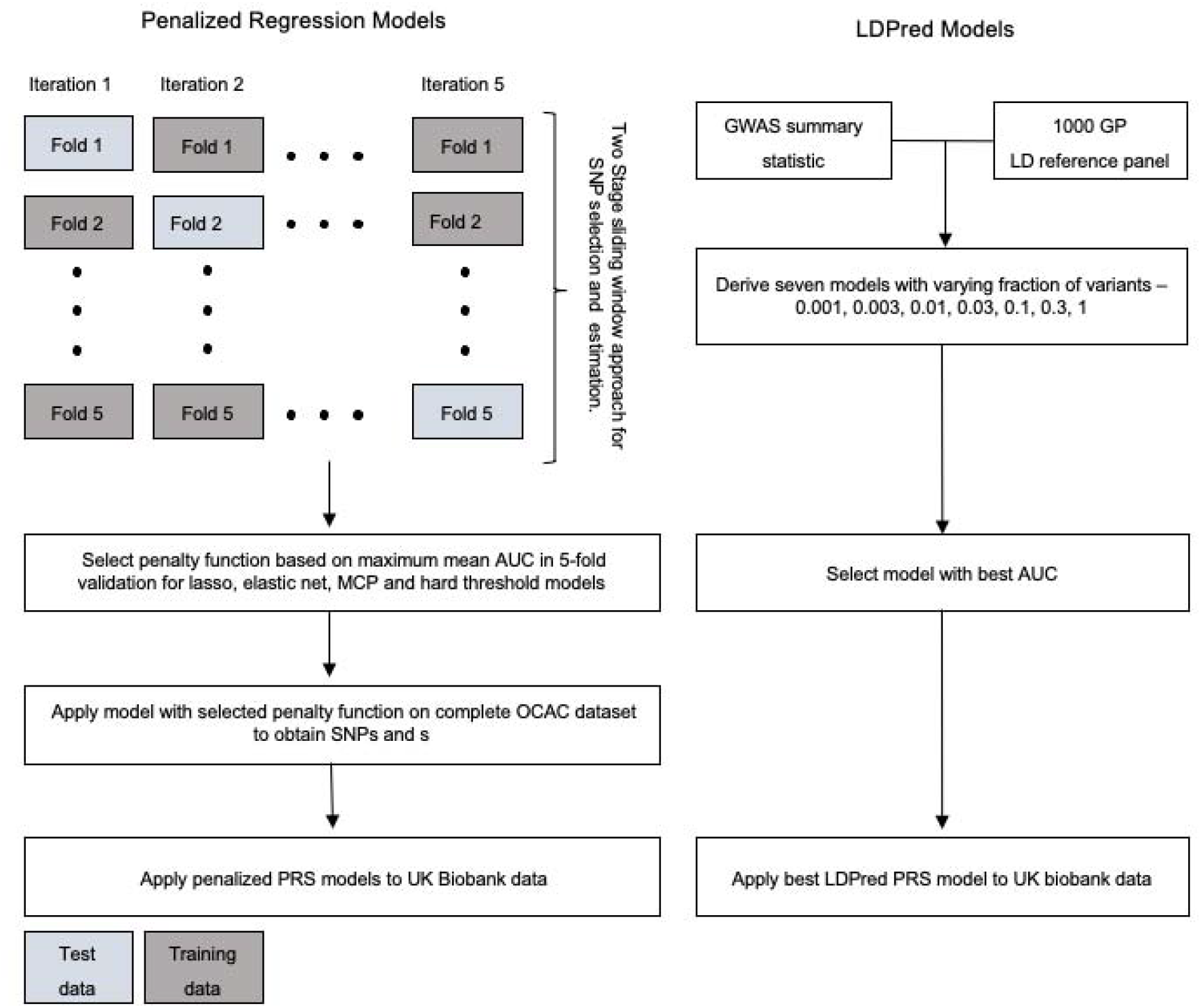
PRS model development using penalized regression and LDPred Bayesian approach.

Finally, we selected the tuning and threshold parameters from the lasso, elastic net and minimax concave penalty models with the maximum mean cross-validated AUC and fitted penalized logistic regression models with these parameters to the entire OCAC dataset to obtain SNP weights for PRS scores.

#### Stepwise logistic regression with variable P-value threshold

This model is a general PLR model with *κ*=1. As with the other PLR models, we investigated various values for *λ* values (corresponding to a variable P-value threshold for including a SNP in the model). However, we observed that the implementation of this model on individual level data was more difficult than for other *κ* values because the model would sometimes converge to a local optimum rather than the global optimum. Therefore, we applied an approximate conditional and joint association analysis using summary level statistics correcting for estimated LD between SNPs, using a reference panel of 5,000 individual level genotype OCAC data as described in Yang et.al (20). Details are provided in the Supplementary Methods.

#### LDPred

LDPred is a Bayesian approach that adjusts GWAS summary statistics for the effects of LD by shrinking the posterior mean effect size of each genetic marker based on a point-normal prior on the effect sizes and LD information from an external reference panel. We derived seven candidate polygenic risk scores assuming the fractions of associated variants were 0.001, 0.003, 0.01, 0.03, 0.1, 0.3 and 1.0 respectively (21) using an LD reference panel of 503 samples of European ancestries from the 1000 Genomes phase 3 release and effect estimates from the genome wide association analysis on the OCAC model development data.

#### Select and shrink using summary statistics (S4)

The S4 algorithm is similar to the PRS-CS algorithm (22) which is a Bayesian approach that uses summary statistics and between-SNP correlation data from a reference panel to generate the PRS scores by placing a continuous shrinkage prior on effect sizes. We adapted this method with penalization of rarer SNPs by correcting for the standard deviation resulting in the selection of fewer SNPs. To implement this algorithm, we varied three parameters, *a, b*, φ, which together control the degree of shrinkage of effect estimates. Φ, the overall shrinkage parameter, is influenced by values of *a* which control shrinkage of effect estimates around 0 and *b* which control shrinkage of larger effect estimates. Smaller values of *a* result in more severe shrinkage of effect estimates than larger values. Conversely, smaller values of *b* produce less severe shrinkage than larger values.

We generated summary statistics for each cross-validation training set and selected the parameters that gave the best results on average from the cross-validation and applied these to the set of summary statistics for the complete OCAC data set to obtain the final set of weights.

#### PRS based on meta-analysis of OCAC-CIMBA summary statistics

We conducted a meta-analysis of the EOC associations in *BRCA1* variant carriers, *BRCA2* variant carriers and the participants participating in OCAC using previously described methodological approaches (16). Additional details are provided in the Supplementary Methods. We constructed two PRS models using results from the OCAC-CIMBA meta-analysis: the Select and Shrink (OCAC-CIMBA) PRS and the Stepwise (OCAC-CIMBA) PRS. To construct the Select and Shrink (OCAC-CIMBA) PRS, we applied the *a, b* and φ parameters from the Select and Shrink model described above to the summary statistics from the meta-analysis to obtain a different set of SNPs and weights. We generated the Stepwise (OCAC-CIMBA) PRS by using histotype-specific results from the meta-analysis. We selected all SNPs that were genome-wide significant at nominal thresholds (p <5×10^−8^), along with any independent signals in the same region with p<10^−5^ from the histotype specific analyses for low-grade serous, high-grade serous, endometrioid, clear cell ovarian cancer and non-mucinous invasive EOC.

#### Polygenic risk score performance

The best lasso, elastic net, stepwise and S4 models from the model development stage were validated using two independent data sources: the UK Biobank data and *BRCA1/BRCA2* pathogenic variant carriers from the CIMBA. In the UK Biobank data, we evaluated discriminatory performance of the models using the AUC and examined the association between standardized PRS and risk of non-mucinous EOC using logistic regression analysis. For the CIMBA data, we assessed associations for each version of the PRS and invasive non-mucinous EOC risk using weighted Cox regression methods previously described (5). PRSs in the CIMBA data were scaled to the same PRS standard deviations as the OCAC data, meaning that per standard deviation hazard ratios estimated on CIMBA data are comparable to PRS associations in the OCAC and UK Biobank data. The regression models were adjusted for birth cohort (<1920, 1920-1929, 1930-1939, 1940-1949, ≥1950) and the first four ancestry informative principal components (calculated separately by iCOGS/OncoArray genotyping array) and stratified by Ashkenazi Jewish ancestry and country. Absolute risks by PRS percentiles adjusting for competing risks of mortality from other causes were calculated as described in the Supplementary Material.

#### Transferability of PRS scores to non-European Ancestry

We implemented two straightforward approaches to disentangle the role of ancestry on polygenic risk scoring. We selected homogenous ancestral samples by using a high cut-off criterion of 80% ancestry and we standardized the polygenic risk scores by mean-centering within each population. These approaches led to a more uniform distribution of polygenic risk scores within each ancestral population. Further adjustments using principal components of ancestry did not attenuate risk estimates.

### Data Availability

OncoArray germline genotype data for the OCAC studies have been deposited at the European Genome-phenome Archive (EGA; https://ega-archive.org/), which is hosted by the EBI and the CRG, under accession EGAS00001002305. Summary results are available from the Ovarian Cancer Association Consortium (http://ocac.ccge.medschl.cam.ac.uk/). A subset of the OncoArray germline genotype data for the CIMBA studies will be made publically available through the database of Genotypes and Phenotypes (dbGaP) under accession phs001321.v1.p1. The complete data set will not be made publically available because of restraints imposed by the ethics committees of individual studies; requests for further data can be made to the Data Access Coordination Committee (http://cimba.ccge.medschl.cam.ac.uk/)

### Ethics Statement

All study participants provided written informed consent and participated in research or clinical studies at the host institute under ethically approved protocols. The studies and their approving institutes are listed in the Supplementary Material (Ethics Statement)

## Results

### Model development

For models based on individual level genotype data, the elastic net model had the best predictive accuracy (model parameters: λ=3.3, *κ*=-2.2, AUC=0.586) Predictive accuracy for the lasso model (λ=3.3, AUC= 0.583) was slightly lower (**Table 1**). The optimal value of λ obtained from regularization paths for the MCP model was 3.3. Further reductions in the degree of penalization for the MCP models did not improve prediction accuracy. Therefore, the best MCP model was equivalent to the lasso model. For models based on summary statistics, the best-fitting S4 model had the best performance (a=2.75, b=2, φ=3e-6, AUC=0.593), whereas the best LDPred model had the poorest performance of the methods tested (p=0.001, AUC=0.552). The mean odds ratios per standard deviation are shown in **Table 1** along with the number of SNPs included in the final model when the models were built with the relevant parameters using the complete dataset. All SNPs selected and the associated weights for each model are provided in Supplementary Tables 1 – 6. Given the poorer performance of the LDpred model and the very large number of SNPs included in the final model it was not considered for further validation in other datasets.

**Table 1:**
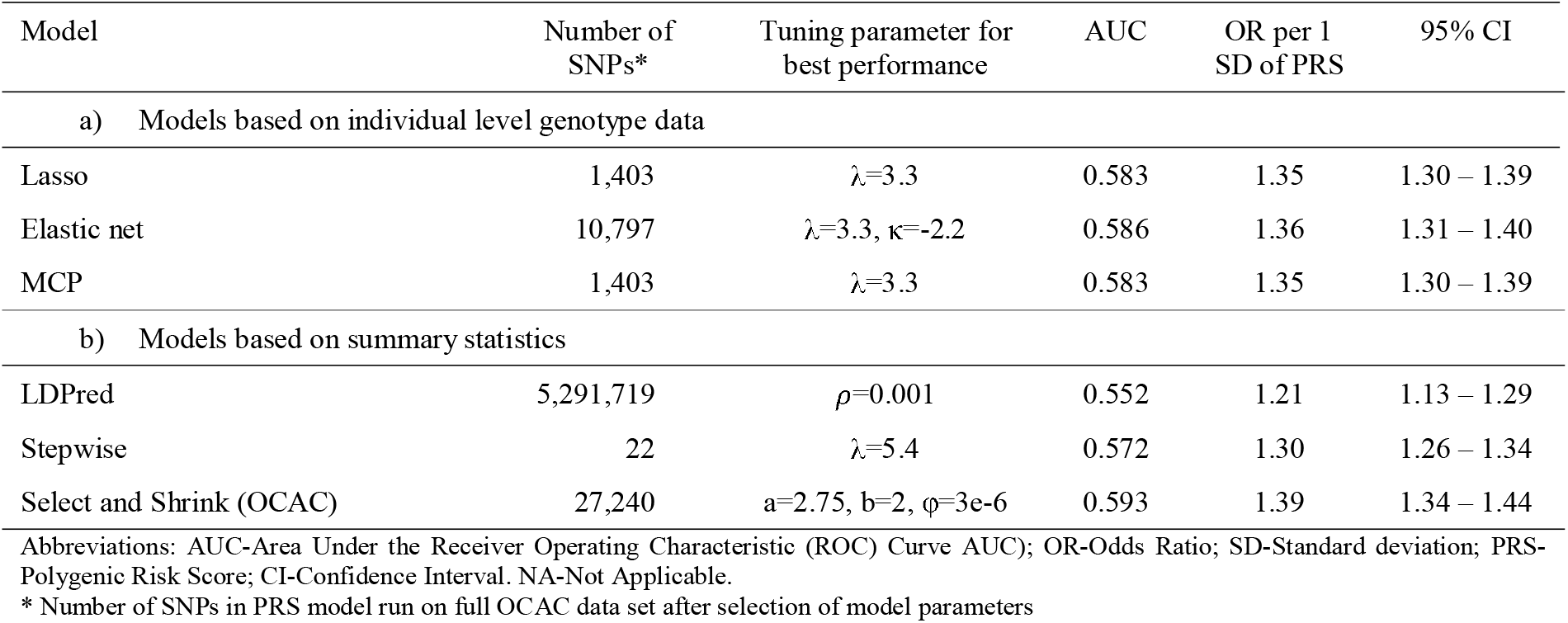
Performance of different PRS models in five-fold cross validation of OCAC data.

### Model validation in women of European ancestry: general population (UK Biobank) and BRCA1/BRCA2 pathogenic variant carriers (CIMBA)

The best AUC estimates derived from cross-validation are likely to be upwardly biased due to overfitting. Therefore, we used the UK Biobank data as an external validation dataset. Overall the PLR models performed slightly better in the UK Biobank data than the model development data (**Table 2**). Of the models developed using the OCAC model development data, the association between PRS and non-mucinous EOC was strongest with the Select and Shrink derived PRS (OR per unit SD=1.38, 95%CI:1.28– 1.48) and slightly lower for the lasso PRS (OR per unit SD=1.37, 95%CI:1.27–1.48), the elastic net PRS (OR per unit SD=1.36, 95%CI:1.26–1.47) and the stepwise PRS model (OR per unit SD=1.35, 95%CI:1.26–1.46). In *BRCA1* and *BRCA2* variant carriers, prediction accuracy was generally higher among *BRCA2* carriers than *BRCA1* carriers. Consistent with results from the general population in the UK Biobank, the Select and Shrink PRS model also had the strongest association and predictive accuracy for invasive EOC risk in both *BRCA1* (HR per unit SD=1.37, 95%CI:1.30–1.44, AUC=0.592) carriers and *BRCA2* carriers (HR per unit SD=1.51, 95%CI:1.36–1.67, AUC=0.624). The PRS models developed using the OCAC-CIMBA meta-analysis results had better discriminative ability in the UK Biobank than the PRS models developed using only OCAC data. Compared with the Select and Shrink model using only OCAC data, the Select and Shrink PRS model derived from the meta-analysis had fewer SNPs (n=18,007), a stronger association with invasive EOC risk (OR per unit SD=1.42, 95%CI:1.32–1.54) and better predictive accuracy (AUC=0.596). Similarly, the Stepwise model from the OCAC-CIMBA meta-analysis performed better than the Stepwise model from only OCAC data (OR per unit SD=1.39, 95%CI:1.29–1.50, AUC=0.595), but included more SNPs (n=36)

**Table 2:**
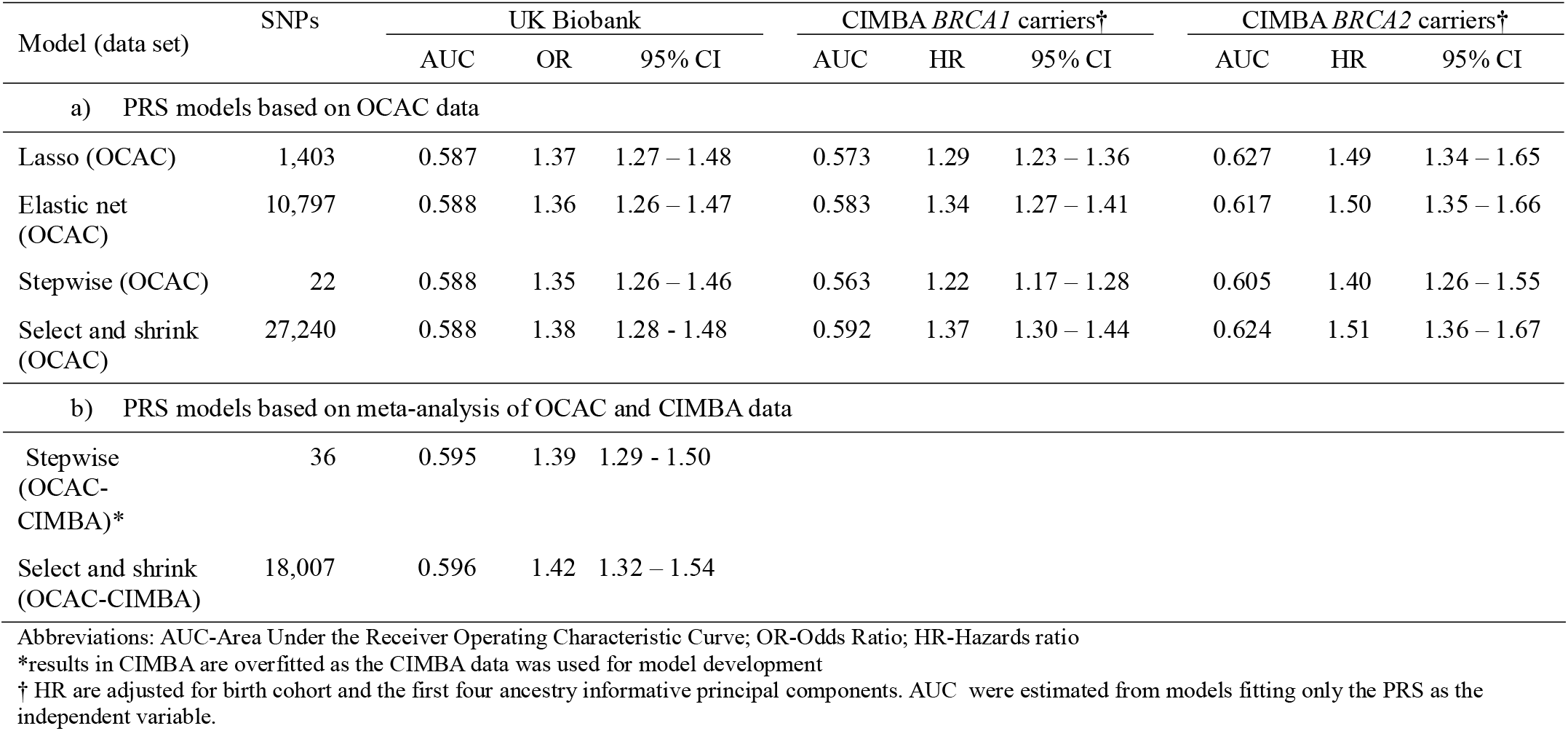
External validation of PRS models in European populations using data from UK Biobank and CIMBA.

Compared with women in the middle quintile, women in the top 95^th^ percentile of the lasso derived PRS model had 2.23-fold increased odds of non-mucinous EOC (**Table 3**). The observed distribution of the OR estimates was consistent with ORs obtained from theoretical predicted values under the assumption that all SNPs interact multiplicatively, especially for the lasso model (**Figure 3**), with all 95% confidence intervals intersecting with the theoretical estimates for women of European ancestry.

**Table 3:**
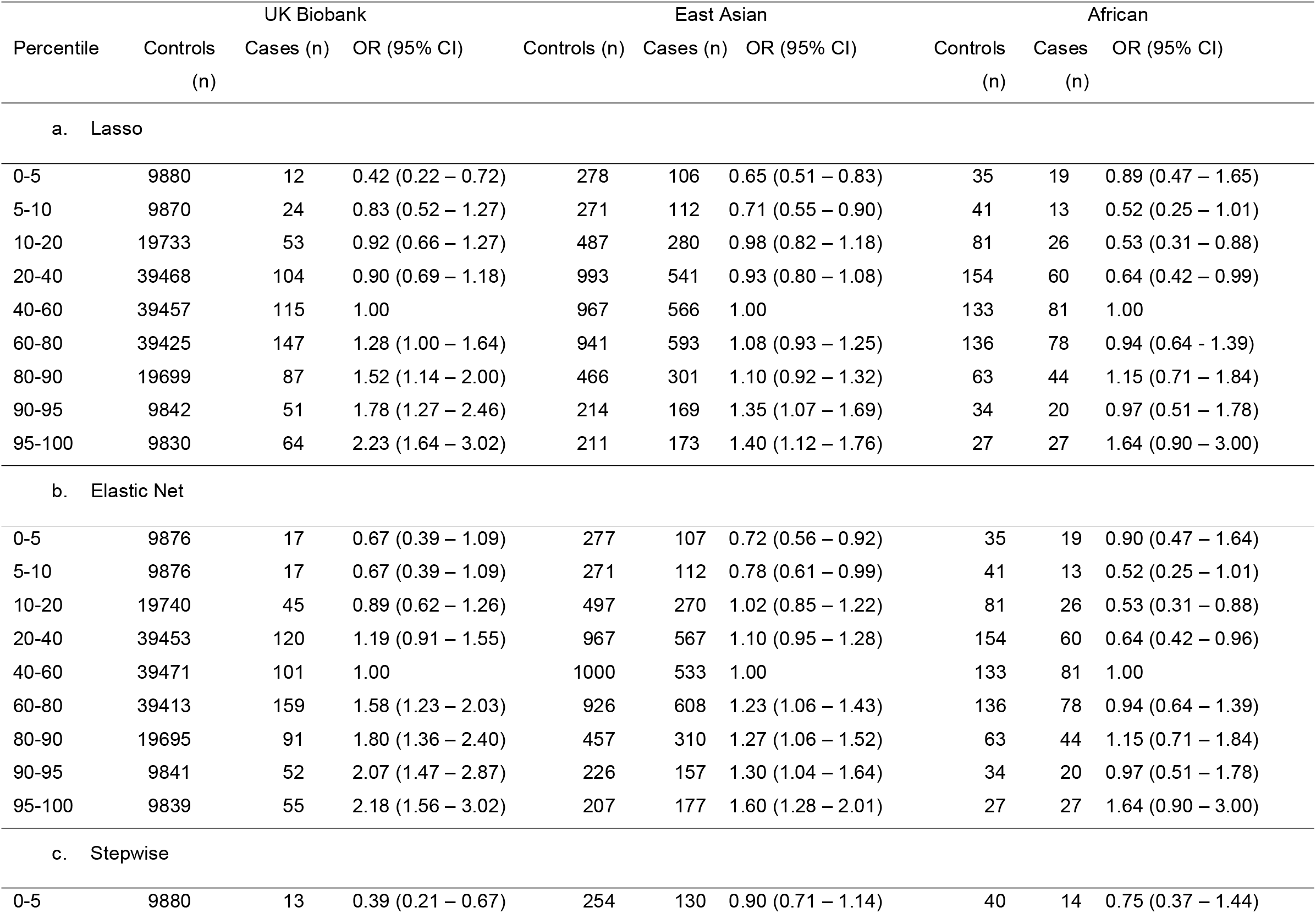
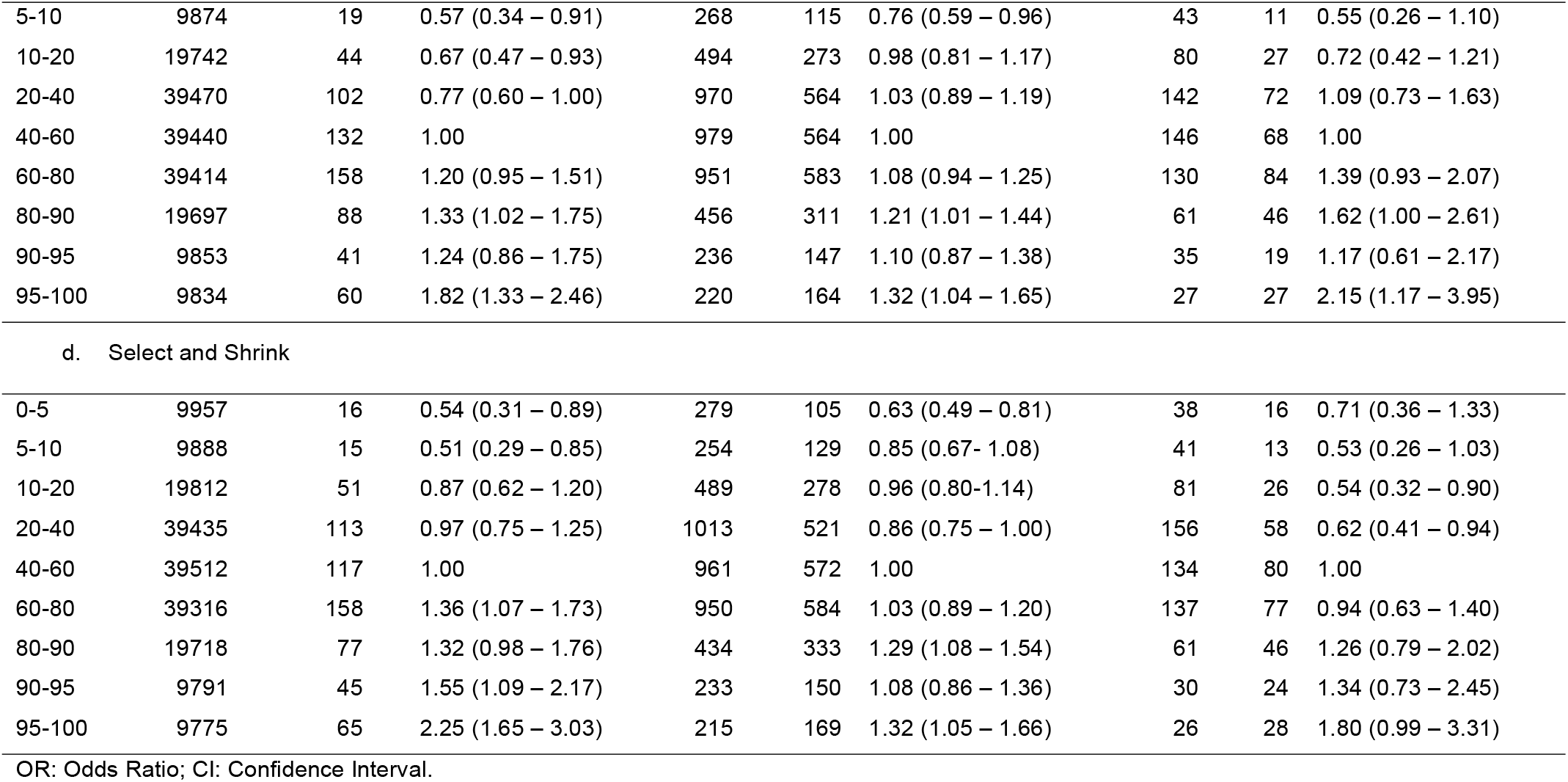
Association between polygenic risk scores and non-mucinous EOC by PRS percentiles and ancestry.

| Percentile | UK Biobank |  |  | East Asian |  |  | African |  |  |
| --- | --- | --- | --- | --- | --- | --- | --- | --- | --- |
|  | Controls (n) | Cases (n) | OR (95% CI) | Controls (n) | Cases (n) | OR (95% CI) | Controls (n) | Cases (n) | OR (95% CI) |
| **a. Lasso** |  |  |  |  |  |  |  |  |  |
| 0-5 | 9880 | 12 | 0.42 (0.22 – 0.72) | 278 | 106 | 0.65 (0.51 – 0.83) | 35 | 19 | 0.89 (0.47 – 1.65) |
| 5-10 | 9870 | 24 | 0.83 (0.52 – 1.27) | 271 | 112 | 0.71 (0.55 – 0.90) | 41 | 13 | 0.52 (0.25 – 1.01) |
| 10-20 | 19733 | 53 | 0.92 (0.66 – 1.27) | 487 | 280 | 0.98 (0.82 – 1.18) | 81 | 26 | 0.53 (0.31 – 0.88) |
| 20-40 | 39468 | 104 | 0.90 (0.69 – 1.18) | 993 | 541 | 0.93 (0.80 – 1.08) | 154 | 60 | 0.64 (0.42 – 0.99) |
| 40-60 | 39457 | 115 | 1.00 | 967 | 566 | 1.00 | 133 | 81 | 1.00 |
| 60-80 | 39425 | 147 | 1.28 (1.00 – 1.64) | 941 | 593 | 1.08 (0.93 – 1.25) | 136 | 78 | 0.94 (0.64 – 1.39) |
| 80-90 | 19699 | 87 | 1.52 (1.14 – 2.00) | 466 | 301 | 1.10 (0.92 – 1.32) | 63 | 44 | 1.15 (0.71 – 1.84) |
| 90-95 | 9842 | 51 | 1.78 (1.27 – 2.46) | 214 | 169 | 1.35 (1.07 – 1.69) | 34 | 20 | 0.97 (0.51 – 1.78) |
| 95-100 | 9830 | 64 | 2.23 (1.64 – 3.02) | 211 | 173 | 1.40 (1.12 – 1.76) | 27 | 27 | 1.64 (0.90 – 3.00) |
| **b. Elastic Net** |  |  |  |  |  |  |  |  |  |
| 0-5 | 9876 | 17 | 0.67 (0.39 – 1.09) | 277 | 107 | 0.72 (0.56 – 0.92) | 35 | 19 | 0.90 (0.47 – 1.64) |
| 5-10 | 9876 | 17 | 0.67 (0.39 – 1.09) | 271 | 112 | 0.78 (0.61 – 0.99) | 41 | 13 | 0.52 (0.25 – 1.01) |
| 10-20 | 19740 | 45 | 0.89 (0.62 – 1.26) | 497 | 270 | 1.02 (0.85 – 1.22) | 81 | 26 | 0.53 (0.31 – 0.88) |
| 20-40 | 39453 | 120 | 1.19 (0.91 – 1.55) | 967 | 567 | 1.10 (0.95 – 1.28) | 154 | 60 | 0.64 (0.42 – 0.96) |
| 40-60 | 39471 | 101 | 1.00 | 1000 | 533 | 1.00 | 133 | 81 | 1.00 |
| 60-80 | 39413 | 159 | 1.58 (1.23 – 2.03) | 926 | 608 | 1.23 (1.06 – 1.43) | 136 | 78 | 0.94 (0.64 – 1.39) |
| 80-90 | 19695 | 91 | 1.80 (1.36 – 2.40) | 457 | 310 | 1.27 (1.06 – 1.52) | 63 | 44 | 1.15 (0.71 – 1.84) |
| 90-95 | 9841 | 52 | 2.07 (1.47 – 2.87) | 226 | 157 | 1.30 (1.04 – 1.64) | 34 | 20 | 0.97 (0.51 – 1.78) |
| 95-100 | 9839 | 55 | 2.18 (1.56 – 3.02) | 207 | 177 | 1.60 (1.28 – 2.01) | 27 | 27 | 1.64 (0.90 – 3.00) |
| **c. Stepwise** |  |  |  |  |  |  |  |  |  |
| 0-5 | 9880 | 13 | 0.39 (0.21 – 0.67) | 254 | 130 | 0.90 (0.71 – 1.14) | 40 | 14 | 0.75 (0.37 – 1.44) |
|  | Controls (n) | Cases (n) | OR (95% CI) | Controls (n) | Cases (n) | OR (95% CI) | Controls (n) | Cases (n) | OR (95% CI) |
| 5-10 | 9874 | 19 | 0.57 (0.34 – 0.91) | 268 | 115 | 0.76 (0.59 – 0.96) | 43 | 11 | 0.55 (0.26 – 1.10) |
| 10-20 | 19742 | 44 | 0.67 (0.47 – 0.93) | 494 | 273 | 0.98 (0.81 – 1.17) | 80 | 27 | 0.72 (0.42 – 1.21) |
| 20-40 | 39470 | 102 | 0.77 (0.60 – 1.00) | 970 | 564 | 1.03 (0.89 – 1.19) | 142 | 72 | 1.09 (0.73 – 1.63) |
| 40-60 | 39440 | 132 | 1.00 | 979 | 564 | 1.00 | 146 | 68 | 1.00 |
| 60-80 | 39414 | 158 | 1.20 (0.95 – 1.51) | 951 | 583 | 1.08 (0.94 – 1.25) | 130 | 84 | 1.39 (0.93 – 2.07) |
| 80-90 | 19697 | 88 | 1.33 (1.02 – 1.75) | 456 | 311 | 1.21 (1.01 – 1.44) | 61 | 46 | 1.62 (1.00 – 2.61) |
| 90-95 | 9853 | 41 | 1.24 (0.86 – 1.75) | 236 | 147 | 1.10 (0.87 – 1.38) | 35 | 19 | 1.17 (0.61 – 2.17) |
| 95-100 | 9834 | 60 | 1.82 (1.33 – 2.46) | 220 | 164 | 1.32 (1.04 – 1.65) | 27 | 27 | 2.15 (1.17 – 3.95) |
| d. Select and Shrink |  |  |  |  |  |  |  |  |  |
| 0-5 | 9957 | 16 | 0.54 (0.31 – 0.89) | 279 | 105 | 0.63 (0.49 – 0.81) | 38 | 16 | 0.71 (0.36 – 1.33) |
| 5-10 | 9888 | 15 | 0.51 (0.29 – 0.85) | 254 | 129 | 0.85 (0.67- 1.08) | 41 | 13 | 0.53 (0.26 – 1.03) |
| 10-20 | 19812 | 51 | 0.87 (0.62 – 1.20) | 489 | 278 | 0.96 (0.80-1.14) | 81 | 26 | 0.54 (0.32 – 0.90) |
| 20-40 | 39435 | 113 | 0.97 (0.75 – 1.25) | 1013 | 521 | 0.86 (0.75 – 1.00) | 156 | 58 | 0.62 (0.41 – 0.94) |
| 40-60 | 39512 | 117 | 1.00 | 961 | 572 | 1.00 | 134 | 80 | 1.00 |
| 60-80 | 39316 | 158 | 1.36 (1.07 – 1.73) | 950 | 584 | 1.03 (0.89 – 1.20) | 137 | 77 | 0.94 (0.63 – 1.40) |
| 80-90 | 19718 | 77 | 1.32 (0.98 – 1.76) | 434 | 333 | 1.29 (1.08 – 1.54) | 61 | 46 | 1.26 (0.79 – 2.02) |
| 90-95 | 9791 | 45 | 1.55 (1.09 – 2.17) | 233 | 150 | 1.08 (0.86 – 1.36) | 30 | 24 | 1.34 (0.73 – 2.45) |
| 95-100 | 9775 | 65 | 2.25 (1.65 – 3.03) | 215 | 169 | 1.32 (1.05 – 1.66) | 26 | 28 | 1.80 (0.99 – 3.31) |

OR: Odds Ratio; CI: Confidence Interval.

### Absolute Risk of Developing Ovarian Cancer by PRS percentiles

We estimated cumulative risk of EOC experienced between birth and the age of 80 within PRS percentiles for women in the general population (**Figure 2**), by applying the odds ratio from the PRS models to age-specific population incidence and mortality data for England in 2016. For *BRCA1* and *BRCA2* pathogenic variant carriers, we applied the estimated hazard ratios from PRS models to age-specific incidence rates obtained from Kuchenbaecker et al. (23).

**Figure 2:**
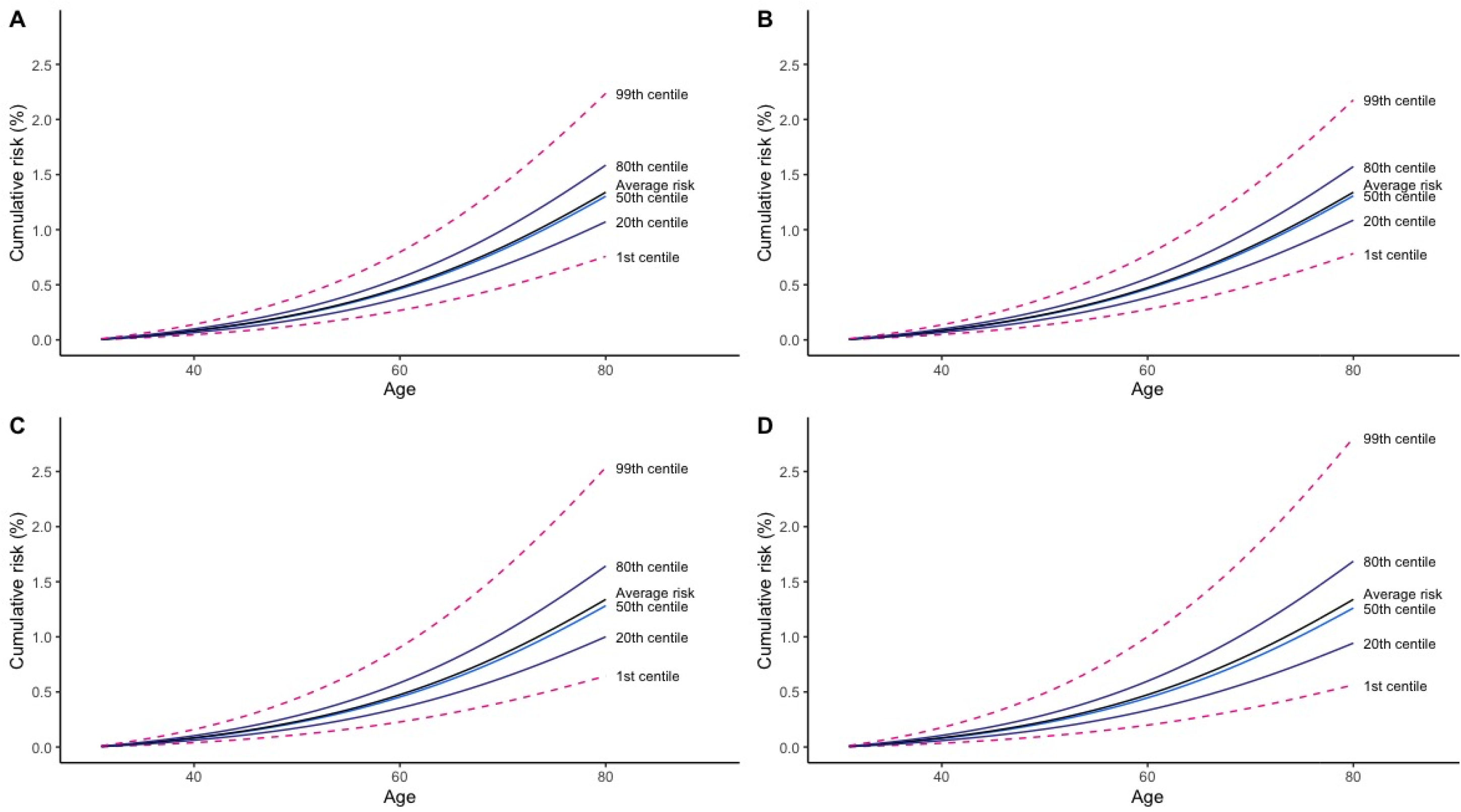
Cumulative risk of ovarian cancer between birth and age 80 by PRS percentiles and PRS models. Shown are the cumulative risk of ovarian cancer risk in UK women by polygenic risk score percentiles. The lasso (A) and elastic net (B) penalized regression models were applied to individual level genotype data, while the stepwise (C) and S4 (D) models were applied to summary level statistics.

**Figure 3:**
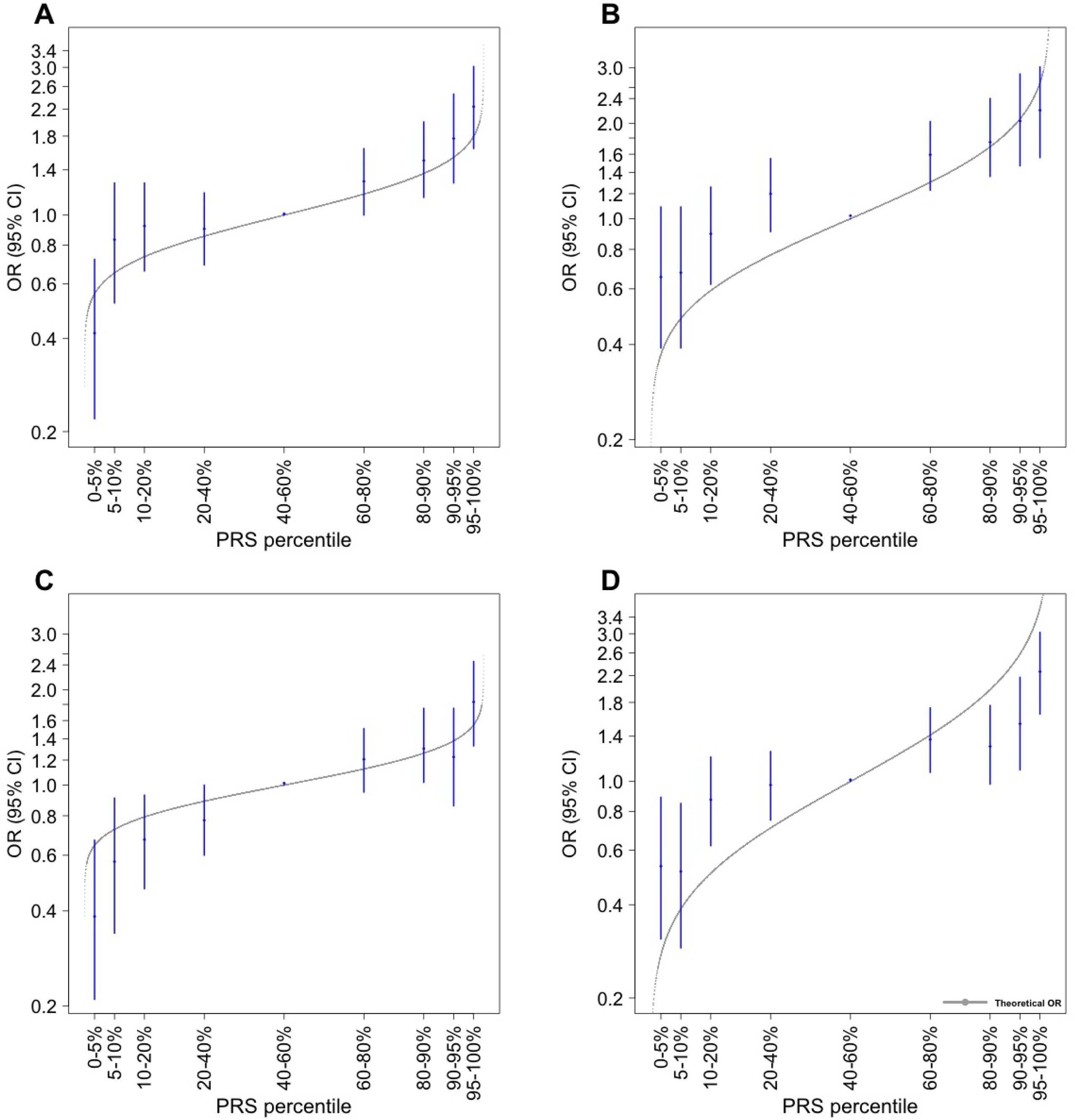
Association between the PLR PRS models and non-mucinous ovarian cancer by PRS percentiles. Shown are estimated odds ratios (OR) and confidence intervals for women of European ancestry by percentiles of polygenic risk scores derived from lasso (A), elastic net (B), stepwise (C) and S4 (D) models relative to the middle quintile.

For women in the general population, the estimated cumulative risks of EOC by age 80 for women at the 99^th^ centile of the PRS distribution were 2.24%, 2.18%, 2.54% and 2.81% for the lasso, elastic net, stepwise and S4 models, respectively. In comparison, the absolute risks of EOC by age 80 for women at the 1^st^ centile were 0.76%, 0.78%, 0.64% and 0.56% for the lasso, elastic net, stepwise and S4 models, respectively.

The absolute risks of developing EOC in *BRCA1* and *BRCA2* pathogenic variant carriers were considerably higher than for women in the general population (Figures S1:Cumulative risk of ovarian cancer risk in *BRCA1* carriers by polygenic risk score percentiles and S2: Cummulative risk of ovarian cancer risk in *BRCA2* carriers by polygenic risk score percentiles). The estimated absolute risk of developing ovarian cancer by age 80 for *BRCA1* carriers at the 99^th^ PRS centiles were 63.2%, 66.3%, 59.0% and 68.4% for the lasso, elastic net, stepwise and S4 models, respectively. The corresponding absolute risks for women at the 1^st^ PRS centile were 27.7%, 25.6%, 30.8% and 24.2%. Absolute risks of developing EOC were lower for *BRCA2* carriers than *BRCA1* carriers, with absolute risks for women in the 99^th^ centile being 36.3%, 36.3%, 33.0% and 36.9%; and absolute risks for women in the 1^st^ centile being 7.10%, 7.12%, 8.24% and 6.92% for the lasso, elastic net, stepwise and S4 models, respectively. Absolute risks for *BRCA1* and *BRCA2* carriers at the 10^th^ and 90^th^ percentile are provided in Supplementary Table 7.

### PRS distribution and ancestry

To investigate the transferability of the PLR derived PRS to other populations, we applied the scores to women of African (N=1,072) and Asian (N=7,669) ancestry genotyped as part of the OncoArray project. In general, the distributions of the raw polygenic scores were dependent on both the statistical methods used in SNP selection and ancestral group. PRS models that included more variants had less dispersion, such that the elastic net models had the least between individual variation in all ancestral groups (standard deviation=0.15, 0.19 and 0.22 for individuals of Asian, African and European ancestries respectively), while the distributions from the stepwise models were the most dispersed (standard deviation = 0.23, 0.27 and 0.30 for individuals of Asian, African and European ancestries respectively). As expected, given the variation in variant frequencies by population, the distribution of polygenic scores was significantly different across the three ancestral groups, with the least dispersion among women of Asian ancestry and the most variation in women of European ancestry. The difference in polygenic risk score distribution was minimized after correction for ancestry by standardizing the PRS to have unit standard deviation using the control subjects for each ancestral group. For comparison, we investigated the use of the first 20 principal components to correct for ancestry and we obtained similar results.

High PRSs were significantly associated with risk of non-mucinous EOC in both Asian and African ancestries (**Table 4**), although the effects were weaker than in women of European ancestry. For example, with the lasso model, the odds ratio (95% CI) per unit standard deviation increment in polygenic score was estimated to be 1.16 (1.11–1.22) in women of

**Table 4:** External validation of PRS models in East Asian and African Populations.

| Model | East Asian ancestries |  |  | African ancestries |  |  |
| --- | --- | --- | --- | --- | --- | --- |
|  | AUC | OR | 95% CI | AUC | OR | 95% CI |
| Lasso | 0.541 | 1.16 | (1.11 – 1.22) | 0.576 | 1.28 | (1.13 – 1.45) |
| Elastic net | 0.543 | 1.17 | (1.12 – 1.23) | 0.574 | 1.29 | (1.14 – 1.47) |
| Stepwise (OCAC) | 0.528 | 1.11 | (1.06 – 1.16) | 0.581 | 1.34 | (1.18 – 1.52) |
| Select and shrink (OCAC) | 0.538 | 1.14 | (1.08 – 1.19) | 0.593 | 1.38 | (1.21 – 1.58) |
| Stepwise<br>(OCAC-CIMBA) | 0.542 | 1.17 | (1.11 – 1.23) | 0.594 | 1.37 | (1.20 – 1.56) |
| Select and shrink<br>(OCAC-CIMBA) | 0.537 | 1.14 | (1.08 – 1.19) | 0.596 | 1.41 | (1.23 – 1.61) |

East Asian ancestry and 1.28 (1.13–1.45) in women of African ancestry compared to 1.37 (1.27–1.48) in women of European ancestry (p for heterogeneity < 0.0001). Variability in effect sizes among ancestral groups was highest for the stepwise model (*I*^2^ = 92%) *versus* 84% and 83% for elastic net and lasso derived polygenic scores respectively. The best discriminative model among women of East Asian and African ancestry were the Elastic net PRS (AUC=0.543) and the Select and Shrink PRS model derived from OCAC-CIMBA meta-analysis (AUC=0.596) respectively. Women of African ancestry in the top 5% of the PRS had about two-fold increased risk compared to women in the middle quintile (lasso OR:1.64,95%CI: 0.90–3.00; elastic net OR:1.64,95%CI:0.90–3.00; stepwise OR:2.15, 95%CI:1.17–3.95; S4 OR:1.80, 95%CI:0.99–3.31). Effect estimates were smaller in women of East Asian ancestry with women in the top 5% of the PRS, having about a 1.5 fold increased risk compared to women in the middle quintile (lasso OR:1.40, 95%CI:1.12–1.76; elastic net OR:1.60, 95% CI:1.28–2.01; stepwise OR:1.32, 95%CI:1.04–1.65; S4 OR:1.32, 95%CI:1.05–1.66).

## Discussion

Genetic risk profiling with polygenic risk scores has led to actionable outcomes for cancers such as breast and prostate (24,25). Previous PRS scores for invasive EOC risk in the general population and *BRCA1/BRCA2* pathogenic variant carriers have been based on genetic variants for which an association with EOC risk had been established at nominal genome-wide significance (4,5,26–28). Here, we explored the predictive performance of computationally-efficient, penalized, regression methods in modelling joint SNP effects for EOC risk prediction in diverse populations and compared them with common approaches. By leveraging the correlation between SNPs which do not reach nominal genome-wide thresholds and including them in PRS models, the polygenic risk scores derived from penalized regression models in this analysis provide stronger evidence of association with risk of non-mucinous EOC than previously published PRSs in both the general population and in *BRCA1/BRCA2* pathogenic variant carriers.

Recently, Barnes et. al derived a PRS score using 22 SNPs that were significantly associated with high-grade serous EOC risk in GWAS (PRS_HGS_) to predict EOC risk in *BRCA1/BRCA2* pathogenic variant carriers (5). To make effect estimates obtained in this analysis comparable to the effect estimates obtained from the PRS_HGS_, we standardized all PRSs using the standard deviation from unaffected *BRCA1/BRCA2* carriers; all PRS models in this analysis except the Stepwise (OCAC only) had higher effect estimates (5). However, the corresponding AUCs were higher for the PRS_HGS_ model (0.604 for *BRCA1* carriers and 0.667 for *BRCA2* carriers), most likely as a result of inclusion of other predictors (birth cohort and principal components) in the model. The AUC estimates for women in the general population, as estimated from the UK Biobank, are slightly higher than estimates from previously published PRS models for overall EOC risk by Wei et al (AUC=0.57) and Yang et al (AUC=0.58) (26,28)

In theory, polygenic risk profiling has the potential for clinical utility, being the earliest measurable contributor to risk which may lead to actionable outcomes. The level of risk among women considered to have a high polygenic risk score, for example women in the 95^th^ percentile, for all of the models we considered approaches the same level of risk conferred by pathogenic variants in moderate penetrance genes such as *FANCM* (RR=2.1, 95%CI=1.1– 3.9) and *PALB2* (RR=2.91 95%CI=1.40–6.04) (29,30). The inclusion of other risk factors such as family history of ovarian cancer, presence of rare pathogenic variants, age at menarche, oral contraceptive use, hormone replacement therapy, parity, and endometriosis in combination with the PRS models could potentially improve risk stratification as has been implemented in the CanRisk tool (www.canrisk.org), which currently uses a PRS model based on 36 SNPs with the potential to use other PRS models (31,32).

An important consideration in the clinical utility of polygenic risk scores is the degree to which results are applicable to diverse populations. We found that the discriminative ability varied substantially by ancestral group. As expected, given that the model development dataset consisted entirely of women of European ancestry, the models had greater discriminative power in women of European ancestry, relative to women of African and East Asian ancestry. We observed greater attenuation of discriminative ability in East Asian populations than African populations. This finding is in contrast to what one would expect given human demographic history, and results from genome wide association studies for EOC (18,19,33,34). One possible explanation for this disparity is the small sample size and imprecise effect estimates for women of African ancestry in this study, due to the larger differences in allele frequency between this population and that of the cohort used to develop the model. Although the model development data for this analysis was predominantly women of European ancestry, the models developed using our approach performed substantially better in women of African ancestry than a PRS model developed by combining 24 published GWAS SNPS associated with non-mucinous EOC, for which the odds of EOC risk was 1.20 fold per standard deviation of PRS (19).

Further refinements to our models, by exploring other penalty functions, may improve the predictive value of the PRS. However, this approach may be complicated by difficulties that arise due to the correlation structure between SNPs. Another option to optimizing the models could be varying the penalization function based on prior knowledge. In genomic regions that are known to have variants associated with EOC, one is more likely to find other risk-associated variants. Therefore, varying the penalty function in these regions such that more SNPs are selected into the model may improve the PRS. Finally, as more functional data become available, modifying penalty functions to incorporate functional data may further improve the PRS. Current approaches for incorporating functional annotation have resulted in only modest gains in prediction accuracy for complex traits such as breast cancer, celiac disease, type 2 diabetes and rheumatoid arthritis, much of which is attributed to the SNPs selected in the models and not the functional annotation (35).

The UK Biobank, our model validation dataset for women in the general population, had a small number of invasive EOC cases with a disproportionately high number of mucinous cases (166 of the 823 invasive EOC cases or ∼20%). Furthermore, cases of the serous histotype could not be classified as either high-grade or low-grade. Therefore, we could not investigate EOC histotype-specific polygenic scores. As the serous histotype is the most common, it is possible that a high-grade serous EOC specific polygenic score may have better predictive value than a non-mucinous polygenic score.

## Conclusion

In conclusion, our results indicate that using the lasso model for individual level genotype data and the S4 model for summary level data in polygenic risk score construction provide an improvement in risk prediction for non-mucinous EOC over more common approaches. Our approach overcomes the computational limitations in the use of penalized methods for large scale genetic data, particularly in the presence of highly-correlated SNPs and the use of cross-validation for parameter estimation is preferred. In practical terms, the polygenic risk score provides sufficient discrimination, particularly for women of European ancestry, to be considered for inclusion in risk prediction and prevention approaches for EOC in the future. Further studies are required to optimize these polygenic risk scores in ancestrally diverse populations and to validate their performance with the inclusion of other genetic and lifestyle risk factors.

## Data Availability

All data are publicly available

## Acknowledgements and Funding

Full acknowledgement and funding details are provided in the Supplementary Material

