## Supplementary Material for "Polygenic Risk Modelling for Prediction of Epithelial Ovarian Cancer Risk"

**Supplementary Method**

#### Penalized logistic regression models: the lasso, elastic net and minimax concave penalty.

A penalized logistic regression model for a set of SNPs aims to identify a set of regression coefficients that minimize the regularized loss function. The aim is to correct the effect estimates towards zero in order to account for prior likelihood. The regularized loss function is given by

$$plr\left( x;\lambda,\kappa\right)=\left\{ \begin{aligned} {x-\lambda sign\left( x \right)}/{\left( 1-\kappa\right) if\left| x \right|<\lambda/{\kappa and \left| \left( x \right) \right|>\lambda}} \\ x if\left| x \right|\geq\frac{\lambda}{\kappa} \\ 0 if\left| \left( x \right) \right|<\lambda\end{aligned} \right.$$

where $x$ is the estimate of SNP effect, $\lambda$ is the tuning parameter and $\kappa$ is the threshold (penalty) for different regularization paths. $\lambda$ and $\kappa$ are parameters that need to be chosen during model development to optimize performance. The lasso, elastic net, minimax concave penalty (MCP), and p-value thresholds are instances of the function with different $\kappa$ values. At $\kappa=0,$ the function is a lasso regularization path and several regression coefficients are shrunk to zero. When $\kappa<0$, the function is an elastic-net solution. The MCP model is obtained when 0 ≤ *κ* ≤ 1. When $\kappa=1$, the function is equivalent to a P-value threshold model.

As the winner’s curse effect can result in inflated estimates for rarer SNPs, it is desirable to penalize rarer SNPs more heavily than common SNPs. We therefore rescaled and normalized the imputed SNP genotypes (coded as number of alternate alleles carried) as follows:

The alternate allele dose, *d_i_*, is first rescaled so that the mean value is zero

$$d_{i}^{rescaled}= d_{i}-mean(d)$$

The rescaled alternate allele dose is then normalized:

$$d_{i}^{normalized}=\frac{d_{i}^{rescaled}}{\sqrt{\left( \sum_{i} p_{i}(1-p_{i} \right))(var\left( d \right)+0.008)/var(d)}}$$

where *p_i_* is the estimated probability of individual *i* being a case based on a simple logistic regression model. This conversion penalizes rarer SNPs and also means that the tuning parameter, *λ*, used in a penalized logistic regression model corresponds closely to a threshold for including a SNP if the SNP z-score for association is greater than *λ*.

There are two general problems with fitting penalized models to high dimensional genetic data: the large number of SNPs which need to be fitted into memory and the correlation between SNPs. We used a two-stage approach to reduce computational burden without a corresponding loss in predictive power. The first stage was a selection stage to reduce the set of SNPs to a manageable number by excluding those unlikely to be included in a final model. We used a sliding windows approach and split the data into 5.5Mb blocks with a 500kb overlap between blocks. The SNP selection analysis was carried out for each block and the selected SNPs collated. Single SNP association analyses were run, and all SNPs with a X^2^ test statistic of less than 2.25 were excluded. Lasso regression models were then run on the remaining SNPs using lambda values of 3.0 and kappa values (penalty score) of 0.0, 0.2, 0.4, 0.6, 0.8 and 1.0. SNPs selected in any of these models were included in subsequent analyses.

In the second stage, we fit penalized regression models to the training dataset with $\lambda$ values from 3.0 to 5.5 in increments of 0.1 iterated over $\kappa$ values from -3.0 to 1 in increments of 0.1. The lasso model ($\kappa=0$) for each value of lambda was fitted first, which has a unique maximum. From the fitted maximum the kappa value is changed, and the model fitted again. This helps to ensure that the final fitted model is close to the global maximum for non-zero values of kappa. For small lambda values the larger kappa values were not analysed as these would not fit the data well and require significant computational resources and time.

We applied this two-stage approach with five-fold cross-validation by splitting the dataset into five groups, and randomly assigning each individual to one of the five groups. We restricted to OCAC studies with at least five controls and five cases, so that each group in the cross-validation analyses would have cases and controls from all studies in the OCAC data (**Figure 1**). In each iteration, four-groups were used as the training set and the fifth group used as the test set. The variants and their weights from the two-stage penalized logistic regression modelling in the training data were used to calculate the area under the receiver operating characteristic curve (AUC) in the test data. We repeated this process for each cross-validation iteration to obtain a mean AUC for each combination of *λ* and *κ*.

Finally, we selected the tuning and threshold parameters from the lasso, elastic net and minimax concave penalty models with the maximum mean cross-validated AUC and fitted penalized logistic regression models with these parameters to the entire OCAC dataset to obtain SNP weights for PRS scores. Details of the workflow are provided in Figure 1.

We developed a custom program written in C++ to implement the penalized logistic regression PLR models using individual level genotype data. This used the coordinate descent algorithm where the loss of function for each variable is optimized using a univariable Newton step while keeping other variables constant to obtain the optimum penalized likelihood function (36).

#### Stepwise logistic regression with variable P-value threshold

This model is a general PLR model with $\kappa$ = 1. As with the other PLR models, we investigated various values for $\lambda$ values (corresponding to a variable P-value threshold for including a SNP in the model). However, we observed that the implementation of this model on individual level data was more difficult than for other $\kappa$ values because the model would sometimes converge to a local optimum rather than the global optimum. Therefore, we applied an approximate conditional and joint association analysis using summary level statistics correcting for estimated LD between SNPs using a reference panel of 5,000 individual level genotype OCAC data as described in Yang et.al (37). In brief, to initiate the model, we identified the most significant SNP at a specified $\lambda$ value threshold, and subsequently investigated all other SNPs across the genome conditional on the SNP already selected. If a new SNP being jointly tested had correlation r^2^ > 0.9, it was left out of the analysis. The SNP with the minimum conditional SNP lower than the cut-off threshold was selected. This process was repeated until no new SNPs could be selected. All selected SNPs were jointly fitted in the model and SNPs with the largest p-value greater than the cut-off threshold were dropped. This process was repeated until no new SNPs were selected or dropped using the training data. As with the PLR models, we investigated several $\lambda$ values. We used the same cross-validated sets as were used by the PLR methods. The lambda coefficient that gave the best average likelihood increase in the cross-validation test set was applied to the full OCAC data set to obtain the final set of weights.

***Meta-analysis of OCAC-CIMBA summary statistics***

We conducted a meta-analysis of the EOC associations in *BRCA1* pathogenic variant carriers, *BRCA2* pathogenic variant carriers and the participants participating in OCAC using previously described methodological approaches (3). In brief, the primary analyses of SNPs and EOC risk in *BRCA1/BRCA2* pathogenic variant carriers included samples genotyped on the iCOGS and OncoArray genotyping chips (with a preference for data from the OncoArray chip when samples were genotyped on both platforms), excluded all overlapping OCAC samples (148 *BRCA1* and 51 *BRCA2*) and SNPs not included in the OncoArray chip. Association analyses were carried out separately for *BRCA1* pathogenic variant carriers and *BRCA2* pathogenic variant carriers, within a survival analysis framework, with time to ovarian cancer diagnosis as the end point. Hazards ratios (HRs) from BRCA1 and BRCA2 pathogenic variants carriers were pooled to give an overall *BRCA1*+*BRCA2* effect estimate. SNPs exhibiting evidence of heterogeneity (P_het_<1x10^-4^) were not considered for downstream analyses. Per-allele HR estimates from *BRCA1*+*BRCA2* pathogenic variant carriers were combined with per allele odds ratios from OCAC to provide an overall relative risk invasive EOC. SNPs with allele frequencies <0.5% were excluded from consideration. All meta-analyses were fixed-effects inverse-variance weighted and implemented using the METAL software (39).

#### Absolute risk calculations

We calculated the absolute risk of EOC by PRS percentiles adjusting for competing risks of mortality from other causes. We obtained incidence rates of EOC associated with an average risk profile and mortality estimates from 2016 population-based registries in the UK. The absolute risk of EOC in PRS percentiles at a given age (AR_prs_ (t)) is calculated as

$${AR}_{PRS}\left( t \right)=\sum_{u=0}^{t} I_{PRS}\left( u \right){.S}_{PRS}\left( u \right).S_{m}\left( u \right)$$

Where $I_{PRS}\left( u \right)$ is the EOC incidence associated with the PRS at age t, $S_{PRS}\left( t \right)$ is the PRS-percentile specific probability of being free of ovarian cancer up to age t and $S_{m}\left( t \right)$ is the probability of surviving to age t, *i.e.,* not dying from a cause other than EOC.

**SUPPLEMENTARY TABLES**

**Table S1**: Effect estimates (weights) of SNPs selected for lasso PRS model.

**Table S2**: Effect estimates (weights) of SNPs selected for elastic net PRS model.

**Table S3**: Effect estimates (weights) of SNPs selected for Stepwise (OCAC) PRS model.

**Table S4**: Effect estimates (weights) of SNPs selected for Select and Shrink (OCAC) PRS model.

**Table S5**: Effect estimates (weights) of SNPs selected for Stepwise (OCAC-CIMBA) PRS model.

**Table S6**: Effect estimates (weights) of SNPs selected for Slect and Shrink (OCAC-CIMBA) PRS model.

**SUPPLEMENTARY FIGURES**

**Figure S1:** Cumulative risk of ovarian cancer risk in *BRCA1* carriers by polygenic risk score percentiles. The lasso (A) and elastic net (B) penalized regression models were applied to individual level genotype data, while the stepwise (C) and S4 (D) models were applied to summary level statistics.

**Figure S2:** Cumulative risk of ovarian cancer risk in *BRCA2* carriers by polygenic risk score percentiles. The lasso (A) and elastic net (B) penalized regression models were applied to individual level genotype data, while the stepwise (C) and S4 (D) models were applied to summary level statistics.

### ETHICS STATEMENT

All study participants provided written informed consent and participated in research studies at the host institute under ethically approved protocols.

**OCAC FUNDING AND ACKNOWLEDGEMENT**

**Funding**

The Ovarian Cancer Association Consortium is supported by a grant from the Ovarian Cancer Research Fund thanks to donations by the family and friends of Kathryn Sladek Smith (PPD/RPCI.07). The scientific development and funding for this project were in part supported by the US National Cancer Institute GAME-ON Post-GWAS Initiative (U19-CA148112). This study made use of data generated by the Wellcome Trust Case Control consortium that was funded by the Wellcome Trust under award 076113. The results published here are in part based upon data generated by The Cancer Genome Atlas Pilot Project established by the National Cancer Institute and National Human Genome Research Institute (dbGap accession number phs000178.v8.p7).

The OCAC OncoArray genotyping project was funded through grants from the U.S. National Institutes of Health (CA1X01HG007491-01 (C.I.A.), U19-CA148112 (T.A.S.), R01-CA149429 (C.M.P.) and R01-CA058598 (M.T.G.); Canadian Institutes of Health Research (MOP-86727 (L.E.K.) and the Ovarian Cancer Research Fund (A.B.). The COGS project was funded through a European Commission's Seventh Framework Programme grant (agreement number 223175 - HEALTH-F2-2009-223175).

Funding for individual studies: **AAS:** National Institutes of Health (RO1-CA142081); **AOV:** The Canadian Institutes for Health Research (MOP-86727); **AUS:** The Australian Ovarian Cancer Study Group was supported by the U.S. Army Medical Research and Materiel Command under DAMD17-01-1-0729, The Cancer Council Tasmania and The Cancer Foundation of Western Australia and the National Health and Medical Research Council of Australia (NHMRC; ID400413 and ID400281). The Australian Ovarian Cancer Study gratefully acknowledges additional support from S. Boldeman, the Agar family, Ovarian Cancer Action (UK), Ovarian Cancer Australia and the Peter MacCallum Foundation; **BAV:** ELAN Funds of the University of Erlangen-Nuremberg; **BEL:** National Kankerplan; **BGS:** Breast Cancer Now, Institute of Cancer Research; **BVU:** Vanderbilt University Medical Center’s BioVU is supported by the 1S10RR025141-01 instrumentation award and Vanderbilt CTSA grant from the National Institutes of Health (NIH)/National Center for Advancing Translational Sciences (NCATS) (ULTR000445); **CAM:** National Institutes of Health Research Cambridge Biomedical Research Centre and Cancer Research UK Cambridge Cancer Centre; **CHA:** Innovative Research Team in University (PCSIRT) in China (IRT1076); **CNI:** Instituto de Salud Carlos III (PI 12/01319); Ministerio de Economía y Competitividad (SAF2012); **DKE:** Ovarian Cancer Research Fund; **DOV:** National Institutes of Health R01-CA112523 and R01-CA87538; **EMC:** Dutch Cancer Society (EMC 2014-6699); **EPC:** The coordination of EPIC is financially supported by the European Commission (DG-SANCO) and the International Agency for Research on Cancer. The national cohorts are supported by Danish Cancer Society (Denmark) (EMC 2014-6699); Ligue Contre le Cancer, Institut Gustave Roussy, Mutuelle Générale de l’Education Nationale, Institut National de la Santé et de la Recherche Médicale (INSERM) (France); German Cancer Aid, German Cancer Research Center (DKFZ), Federal Ministry of Education and Research (BMBF) (Germany); the Hellenic Health Foundation (Greece); Associazione Italiana per la Ricerca sul Cancro-AIRC-Italy and National Research Council (Italy); Dutch Ministry of Public Health, Welfare and Sports (VWS), Netherlands Cancer Registry (NKR), LK Research Funds, Dutch Prevention Funds, Dutch ZON (Zorg Onderzoek Nederland), World Cancer Research Fund (WCRF), Statistics Netherlands (The Netherlands); ERC-2009-AdG 232997 and Nordforsk, Nordic Centre of Excellence programme on Food, Nutrition and Health (Norway); Health Research Fund (FIS), PI13/00061 to Granada, PI13/01162 to EPIC-Murcia, Regional Governments of Andalucía, Asturias, Basque Country, Murcia and Navarra, ISCIII RETIC (RD06/0020) (Spain); Swedish Cancer Society, Swedish Research Council and County Councils of Skåne and Västerbotten (Sweden); Cancer Research UK (14136 to EPIC-Norfolk; C570/A16491 and C8221/A19170 to EPIC-Oxford), Medical Research Council (1000143 to EPIC-Norfolk, MR/M012190/1 to EPIC-Oxford) (United Kingdom); **GER:** German Federal Ministry of Education and Research, Programme of Clinical Biomedical Research (01 GB 9401) and the German Cancer Research Center (DKFZ); **GRC:** This research has been co-financed by the European Union (European Social Fund - ESF) and Greek national funds through the Operational Program "Education and Lifelong Learning" of the National Strategic Reference Framework (NSRF) - Research Funding Program of the General Secretariat for Research & Technology: SYN11_10_19 NBCA. Investing in knowledge society through the European Social Fund; **GRR:** Roswell Park Cancer Institute Alliance Foundation, P30 CA016056; **HAW:** U.S. National Institutes of Health (R01-CA58598, N01-CN-55424 and N01-PC-67001); **HJO:** Intramural funding; Rudolf-Bartling Foundation; **HMO:** Intramural funding; Rudolf-Bartling Foundation; **HOC:** Helsinki University Hospital Research Fund; **HOP:** University of Pittsburgh School of Medicine Dean’s Faculty Advancement Award (F. Modugno), Department of Defense (DAMD17-02-1-0669) and NCI (K07-CA080668, R01-CA95023, P50-CA159981 MO1-RR000056 R01-CA126841); **HUO:** Intramural funding; Rudolf-Bartling Foundation; **JPN:** Grant-in-Aid for the Third Term Comprehensive 10-Year Strategy for Cancer Control from the Ministry of Health, Labour and Welfare; **KRA:** This study (Ko-EVE) was supported by a grant from the Korea Health Technology R&D Project through the Korea Health Industry Development Institute (KHIDI), and the National R&D Program for Cancer Control, Ministry of Health & Welfare, Republic of Korea (HI16C1127; 0920010); **LAX:** American Cancer Society Early Detection Professorship (SIOP-06-258-01-COUN) and the National Center for Advancing Translational Sciences (NCATS), Grant UL1TR000124; **LUN:** ERC-2011-AdG 294576-risk factors cancer, Swedish Cancer Society, Swedish Research Council, Beta Kamprad Foundation; **MAC:** National Institutes of Health (R01-CA122443, P30-CA15083, P50-CA136393); Mayo Foundation; Minnesota Ovarian Cancer Alliance; Fred C. and Katherine B. Andersen Foundation; Fraternal Order of Eagles; **MAL:** Funding for this study was provided by research grant R01- CA61107 from the National Cancer Institute, Bethesda, MD, research grant 94 222 52 from the Danish Cancer Society, Copenhagen, Denmark; and the Mermaid I project; **MAS:** Malaysian Ministry of Higher Education (UM.C/HlR/MOHE/06) and Cancer Research Initiatives Foundation; **MAY:** National Institutes of Health (R01-CA122443, P30-CA15083, P50-CA136393); Mayo Foundation; Minnesota Ovarian Cancer Alliance; Fred C. and Katherine B. Andersen Foundation; **MCC:** MCCS cohort recruitment was funded by VicHealth and Cancer Council Victoria. Cancer Council Victoria, National Health and Medical Research Council of Australia (NHMRC) grants number 209057, 251533, 396414, and 504715; **MDA:** DOD Ovarian Cancer Research Program (W81XWH-07-0449); **MEC:** NIH (CA54281, CA164973, CA63464); **MOF:** Moffitt Cancer Center, Merck Pharmaceuticals, the state of Florida, Hillsborough County, and the city of Tampa; **NCO:** National Institutes of Health (R01-CA76016) and the Department of Defense (DAMD17-02-1-0666); **NEC:** National Institutes of Health R01-CA54419 and P50-CA105009 and Department of Defense W81XWH-10-1-02802; **NHS:** UM1 CA186107, P01 CA87969, R01 CA49449, R01-CA67262, UM1 CA176726; **NJO:** National Cancer Institute (NIH-K07 CA095666, R01-CA83918, NIH-K22-CA138563, and P30-CA072720) and the Cancer Institute of New Jersey; If Sara Olson and/or Irene Orlow is a co-author, please add NCI CCSG award (P30-CA008748) to the funding sources; **NOR:** Helse Vest, The Norwegian Cancer Society, The Research Council of Norway; **NTH:** Radboud University Medical Centre; **OPL:** National Health and Medical Research Council (NHMRC) of Australia (APP1025142) and Brisbane Women's Club; **ORE:** Sherie Hildreth Ovarian Cancer (SHOC) Foundation; **OVA:** This work was supported by Canadian Institutes of Health Research grant (MOP-86727) and by NIH/NCI 1 R01CA160669-01A1; **PLC:** Intramural Research Program of the National Cancer Institute; **POC:** Pomeranian Medical University; **POL:** Intramural Research Program of the National Cancer Institute; **PVD:** Canadian Cancer Society and Cancer Research Society GRePEC Program; **RBH:** National Health and Medical Research Council of Australia; **RMH:** Cancer Research UK, Royal Marsden Hospital; **RPC:** National Institute of Health (P50 CA159981, R01CA126841); **SEA:** Cancer Research UK (C490/A10119 C490/A10124); UK National Institute for Health Research Biomedical Research Centres at the University of Cambridge; **SIS:** The Sister Study (SISTER) is supported by the Intramural Research Program of the NIH, National Institute of Environmental Health Sciences (Z01-ES044005 and Z01-ES049033); **SMC:** The Swedish Cancer Foundation and the Swedish Research Council (VR 2017-00644) grant for the Swedish Infrastructure for Medical Population-based Life-course Environmental Research (SIMPLER); **SRO:** Cancer Research UK (C536/A13086, C536/A6689) and Imperial Experimental Cancer Research Centre (C1312/A15589); **STA:** NIH grants U01 CA71966 and U01 CA69417; **SWH:** NIH (NCI) grant R37-CA070867; **TBO:** National Institutes of Health (R01-CA106414-A2), American Cancer Society (CRTG-00-196-01-CCE), Department of Defense (DAMD17-98-1-8659), Celma Mastry Ovarian Cancer Foundation; **TOR:** NIH grants R01 CA063678 and R01 CA063682; **UCI:** NIH R01-CA058860 and the Lon V Smith Foundation grant LVS-39420; **UHN:** Princess Margaret Cancer Centre Foundation-Bridge for the Cure; **UKO:** The UKOPS study was funded by The Eve Appeal (The Oak Foundation) with investigators supported by the National Institute for Health Research University College London Hospitals Biomedical Research Centre and MRC core funding (MR_UU_12023); **UKR:** Cancer Research UK (C490/A6187), UK National Institute for Health Research Biomedical Research Centres at the University of Cambridge; **USC:** P01CA17054, P30CA14089, R01CA61132, N01PC67010, R03CA113148, R03CA115195, N01CN025403, and California Cancer Research Program (00-01389V-20170, 2II0200); **VAN:** BC Cancer Foundation, VGH & UBC Hospital Foundation; **VTL:** NIH K05-CA154337; **WMH:** National Health and Medical Research Council of Australia, Enabling Grants ID 310670 & ID 628903. Cancer Institute NSW Grants 12/RIG/1-17 & 15/RIG/1-16; **WOC:** National Science Centre (N N301 5645 40), The Maria Sklodowska-Curie Memorial Cancer Centre and Institute of Oncology, Warsaw, Poland.

This work was supported by the Cedars-Sinai Precision Health Initiative (awarded to Simon Gayther and Michelle Jones*.*

Additional research funding and support included: UM1 CA186107, P01 CA87969, (Nurses’ Health Study); U01 CA176726, R01 CA67262, (Nurses’ Health Study II).

**Acknowledgements**

We are grateful to the family and friends of Kathryn Sladek Smith for their generous support of the Ovarian Cancer Association Consortium through their donations to the Ovarian Cancer Research Fund. The OncoArray and COGS genotyping projects would not have been possible without the contributions of the following: Per Hall (COGS); Douglas F. Easton, Paul Pharoah, Kyriaki Michailidou, Manjeet K. Bolla, Qin Wang (BCAC), Andrew Berchuck, Marjorie J. Riggan (OCAC), Rosalind A. Eeles, Douglas F. Easton, Ali Amin Al Olama, Zsofia Kote-Jarai, Sara Benlloch (PRACTICAL), Georgia Chenevix-Trench, Antonis Antoniou, Lesley McGuffog, Fergus Couch and Ken Offit (CIMBA), Joe Dennis, Jonathan P. Tyrer, Siddhartha Kar, Alison M. Dunning, Andrew Lee, and Ed Dicks, Craig Luccarini and the staff of the Centre for Genetic Epidemiology Laboratory, Javier Benitez, Anna Gonzalez-Neira and the staff of the CNIO genotyping unit, Jacques Simard and Daniel C. Tessier, Francois Bacot, Daniel Vincent, Sylvie LaBoissière and Frederic Robidoux and the staff of the McGill University and Génome Québec Innovation Centre, Stig E. Bojesen, Sune F. Nielsen, Borge G. Nordestgaard, and the staff of the Copenhagen DNA laboratory, and Julie M. Cunningham, Sharon A. Windebank, Christopher A. Hilker, Jeffrey Meyer and the staff of Mayo Clinic Genotyping Core Facility. We pay special tribute to the contribution of Professor Brian Henderson to the GAME-ON consortium; to Olga M. Sinilnikova for her contribution to CIMBA and for her part in the initiation and coordination of GEMO until she sadly passed away on the 30th June 2014 and to Catherine M. Phelan for her contribution to OCAC and coordination of the OncoArray until she passed away on 22 September 2017. We thank the study participants, doctors, nurses, clinical and scientific collaborators, health care providers and health information sources who have contributed to the many studies contributing to this manuscript.

Acknowledgements for individual studies: **AOV:** We thank Jennifer Koziak, Mie Konno, Michelle Darago, Faye Chambers and the Tom Baker Cancer Centre Translational Laboratories; **AUS:** The AOCS also acknowledges the cooperation of the participating institutions in Australia, and the contribution of the study nurses, research assistants and all clinical and scientific collaborators. The complete AOCS Study Group can be found at [www.aocstudy.org](http://www.aocstudy.org). We would like to thank all of the women who participated in this research program; **BEL:** We would like to thank Gilian Peuteman, Thomas Van Brussel, Annick Van den Broeck and Joke De Roover for technical assistance; **BGS:** The Institute of Cancer Research (ICR) acknowledges NHS funding to the NIHR Biomedical Research Centre. We thank the Study staff, study participants , doctors, nurses, health care providers and health information sources who have contributed to the study and Breast Cancer Now and The Institute of Cancer Research for their support; **BVU:** The dataset(s) used for the analyses described were obtained from Vanderbilt University Medical Center’s BioVU; **CHA:** Innovative Research Team in University (PCSIRT) in China (IRT1076); **CHN:** To thank all members of Department of Obstetrics and Gynaecology, Hebei Medical University, Fourth Hospital and Department of Molecular Biology, Hebei Medical University, Fourth Hospital; **COE:** Gynecologic Cancer Center of Excellence; **EPC:** To thank all members and investigators of the Rotterdam Ovarian Cancer Study; **GER:** The German Ovarian Cancer Study (GER) thank Ursula Eilber for competent technical assistance; **MAS:** We would like to thank Famida Zulkifli and Ms Moey for assistance in patient recruitment, data collection and sample preparation; **MCC:** Cases and their vital status were ascertained through the Victorian Cancer Registry (VCR) and the Australian Institute of Health and Welfare (AIHW), including the National Death Index and the Australian Cancer Database; **MOF:** the Total Cancer Care™ Protocol and the Collaborative Data Services and Tissue Core Facilities at the H. Lee Moffitt Cancer Center & Research Institute, an NCI designated Comprehensive Cancer Center (P30-CA076292), Merck Pharmaceuticals and the state of Florida; **NHS:** The NHS/NHSII studies thank the following state cancer registries for their help: AL, AZ, AR, CA, CO, CT, DE, FL, GA, ID, IL, IN, IA, KY, LA, ME, MD, MA, MI, NE, NH, NJ, NY, NC, ND, OH, OK, OR, PA, RI, SC, TN, TX, VA, WA, and WY; **OPL:** Members of the OPAL Study Group (<http://opalstudy.qimrberghofer.edu.au/>); **SEA:** SEARCH team, Craig Luccarini, Caroline Baynes, Don Conroy; **SRO:** To thank all members of Scottish Gynaecological Clinical Trails group and SCOTROC1 investigators; **SWH:** We thank the participants and the research staff of the Shanghai Women’s Health Study for making this study possible; **UHN:** Princess Margaret Cancer Centre Foundation-Bridge for the Cure; **UKO:** We particularly thank I. Jacobs, M.Widschwendter, E. Wozniak, A. Ryan, J. Ford and N. Balogun for their contribution to the study**; UKR:** Carole Pye; **VAN:** BC Cancer Foundation, VGH & UBC Hospital Foundation; **WMH:** We thank the Gynaecological Oncology Biobank at Westmead, a member of the Australasian Biospecimen Network-Oncology group.

The authors would like to thank the participants and staff of the NHS/NHSII for their valuable contributions as well as the following state cancer registries for their help: AL, AZ, AR, CA, CO, CT, DE, FL, GA, ID, IL, IN, IA, KY, LA, ME, MD, MA, MI, NE, NH, NJ, NY, NC, ND, OH, OK, OR, PA, RI, SC, TN, TX, VA, WA, WY. The authors assume full responsibility for analyses and interpretation of these data.

**CIMBA FUNDING AND ACKNOWLEDGEMENT**

**Funding**

The CIMBA data management and data analysis were supported by Cancer Research – UK grants C12292/A20861, C12292/A11174. GCT and ABS are NHMRC Research Fellows. iCOGS: the European Community's Seventh Framework Programme under grant agreement n° 223175 (HEALTH-F2-2009-223175) (COGS), Cancer Research UK (C1287/A10118, C1287/A 10710, C12292/A11174, C1281/A12014, C5047/A8384, C5047/A15007, C5047/A10692, C8197/A16565), the National Institutes of Health (CA128978) and Post- Cancer GWAS initiative (1U19 CA148537, 1U19 CA148065 and 1U19 CA148112 - the GAME-ON initiative), the Department of Defence (W81XWH-10-1-0341), the Canadian Institutes of Health Research (CIHR) for the CIHR Team in Familial Risks of Breast Cancer (CRN-87521), and the Ministry of Economic Development, Innovation and Export Trade (PSR-SIIRI-701), Komen Foundation for the Cure, the Breast Cancer Research Foundation, and the Ovarian Cancer Research Fund. The PERSPECTIVE and PERSPECTIVE I&I projects were supported by the Government of Canada through Genome Canada and the Canadian Institutes of Health Research, the Ministry of Economy and Innovation through Genome Québec, and The Quebec Breast Cancer Foundation and the Ontario Research Fund. BCFR: UM1 CA164920 from the National Cancer Institute. The content of this manuscript does not necessarily reflect the views or policies of the National Cancer Institute or any of the collaborating centers in the Breast Cancer Family Registry (BCFR), nor does mention of trade names, commercial products, or organizations imply endorsement by the US Government or the BCFR. BFBOCC: Lithuania (BFBOCC-LT): Research Council of Lithuania grant SEN-18/2015. BIDMC: Breast Cancer Research Foundation. BMBSA: Cancer Association of South Africa (PI Elizabeth J. van Rensburg). CNIO: Spanish Ministry of Health PI16/00440 supported by FEDER funds, the Spanish Instituto de Salud Carlos III (grant PI19/00640) supported by FEDAR funds and the Spanish Research Network on Rare diseases (CIBERER). COH-CCGCRN: Research reported in this publication was supported by the National Cancer Institute of the National Institutes of Health under grant number R25CA112486, and RC4CA153828 (PI: J. Weitzel) from the National Cancer Institute and the Office of the Director, National Institutes of Health. The content is solely the responsibility of the authors and does not necessarily represent the official views of the National Institutes of Health. CONSIT TEAM: Associazione Italiana Ricerca sul Cancro (AIRC; IG2014 no.15547) to P. Radice. Funds from Italian citizens who allocated the 5x1000 share of their tax payment in support of the Fondazione IRCCS Istituto Nazionale Tumori, according to Italian laws (INT-Institutional strategic projects ‘5x1000’) to S. Manoukian. Associazione Italiana Ricerca sul Cancro (AIRC; IG2015 no.16732) to P. Peterlongo. DEMOKRITOS: European Union (European Social Fund – ESF) and Greek national funds through the Operational Program "Education and Lifelong Learning" of the National Strategic Reference Framework (NSRF) - Research Funding Program of the General Secretariat for Research & Technology: SYN11_10_19 NBCA. Investing in knowledge society through the European Social Fund. DFKZ: German Cancer Research Center. EMBRACE: Cancer Research UK Grants C1287/A17523, C1287/A26886 and C1287/A23382. D. Gareth Evans is supported by an NIHR grant to the Biomedical Research Centre, Manchester (NIHR grant IS-BRC- 1215-20007). The Investigators at The Institute of Cancer Research and The Royal Marsden NHS Foundation Trust are supported by an NIHR grant to the Biomedical Research Centre at The Institute of Cancer Research and The Royal Marsden NHS Foundation Trust. Ros Eeles and Elizabeth Bancroft are supported by Cancer Research UK Grant C5047/A8385. Ros Eeles is also supported by NIHR support to the Biomedical Research Centre at The Institute of Cancer Research and The Royal Marsden NHS Foundation Trust. FCCC: The University of Kansas Cancer Center (P30 CA168524), the Kansas Institute for Precision Medicine COBRE (P20 GM130423), the Kansas Bioscience Authority Eminent Scholar Program, R01 CA140323, R01 CA214545, and by the Chancellors Distinguished Chair in Biomedical Sciences Professorship to A.K.G. Ana Vega is supported by the Spanish Health Research Foundation, Instituto de Salud Carlos III (ISCIII) through Research Activity Intensification Program (contract grant numbers: INT15/00070, INT16/00154, INT17/00133), and through Centro de Investigación Biomédica en Red de Enferemdades Raras CIBERER (ACCI 2016: ER17P1AC7112/2018); Autonomous Government of Galicia (Consolidation and structuring program: IN607B), and by the Fundación Mutua Madrileña (call 2018). GC-HBOC: German Cancer Aid (grant no 110837, Rita K. Schmutzler) and the European Regional Development Fund and Free State of Saxony, Germany (LIFE - Leipzig Research Centre for Civilization Diseases, project numbers 713-241202, 713-241202, 14505/2470, 14575/2470). GEMO: Ligue Nationale Contre le Cancer; the Association “Le cancer du sein, parlons-en!” Award, the Canadian Institutes of Health Research for the "CIHR Team in Familial Risks of Breast Cancer" program and the French National Institute of Cancer (INCa grants 2013-1-BCB-01-ICH-1 and SHS-E-SP 18-015). GEORGETOWN: the Non-Therapeutic Subject Registry Shared Resource at Georgetown University (NIH/NCI grant P30-CA051008), the Fisher Center for Hereditary Cancer and Clinical Genomics Research, and Swing Fore the Cure. G-FAST: Bruce Poppe is a senior clinical investigator of FWO. Mattias Van Heetvelde obtained funding from IWT. HCSC: Spanish Ministry of Health PI15/00059, PI16/01292, and CB- 161200301 CIBERONC from ISCIII (Spain), partially supported by European Regional Development FEDER funds. HEBCS: Helsinki University Hospital Research Fund, the Finnish Cancer Society and the Sigrid Juselius Foundation. The HEBON study is supported by the Dutch Cancer Society grants NKI1998-1854, NKI2004-3088, NKI2007-3756, the Netherlands Organisation of Scientific Research grant NWO 91109024, the Pink Ribbon grants 110005 and 2014-187.WO76, the BBMRI grant NWO 184.021.007/CP46 and the Transcan grant JTC 2012 Cancer 12-054. HRBCP: Hong Kong Sanatorium and Hospital, Dr Ellen Li Charitable Foundation, The Kerry Group Kuok Foundation, National Institute of Health1R 03CA130065, and North California Cancer Center. HUNBOCS: Hungarian Research Grants KTIA-OTKA CK-80745 and NKFI_OTKA K-112228. ICO: The authors would like to particularly acknowledge the support of the Asociación Española Contra el Cáncer (AECC), the Instituto de Salud Carlos III (organismo adscrito al Ministerio de Economía y Competitividad) and “Fondo Europeo de Desarrollo Regional (FEDER), unamanera de hacer Europa” (PI10/01422, PI13/00285, PIE13/00022, PI15/00854, PI16/00563 and CIBERONC) and the Institut Català de la Salut and Autonomous Government of Catalonia (2009SGR290, 2014SGR338 and PERIS Project MedPerCan). IHCC: PBZ_KBN_122/P05/2004. ILUH: Icelandic Association “Walking for Breast Cancer Research” and by the Landspitali University Hospital Research Fund. INHERIT: Canadian Institutes of Health Research for the “CIHR Team in Familial Risks of Breast Cancer” program – grant # CRN-87521 and the Ministry of Economic Development, Innovation and Export Trade – grant # PSR-SIIRI-701. IOVHBOCS: Ministero della Salute and “5x1000” Istituto Oncologico Veneto grant. IPOBCS: Liga Portuguesa Contra o Cancro. kConFab: The National Breast Cancer Foundation, and previously by the National Health and Medical Research Council (NHMRC), the Queensland Cancer Fund, the Cancer Councils of New South Wales, Victoria, Tasmania and South Australia, and the Cancer Foundation of Western Australia. KOHBRA: the Korea Health Technology R&D Project through the Korea Health Industry Development Institute (KHIDI), and the National R&D Program for Cancer Control, Ministry of Health & Welfare, Republic of Korea (HI16C1127; 1020350; 1420190). MAYO: NIH grants CA116167, CA192393 and CA176785, an NCI Specialized Program of Research Excellence (SPORE) in Breast Cancer (CA116201), and a grant from the Breast Cancer Research Foundation. MCGILL: Jewish General Hospital Weekend to End Breast Cancer, Quebec Ministry of Economic Development, Innovation and Export Trade. Marc Tischkowitz is supported by the funded by the European Union Seventh Framework Program (2007Y2013)/European Research Council (Grant No. 310018). MODSQUAD: MH CZ - DRO (MMCI, 00209805), MEYS - NPS I - LO1413 to LF, and by Charles University in Prague project UNCE204024 (MZ). MSKCC: the Breast Cancer Research Foundation, the Robert and Kate Niehaus Clinical Cancer Genetics Initiative, the Andrew Sabin Research Fund and a Cancer Center Support Grant/Core Grant (P30 CA008748). NAROD: 1R01 CA149429-01. NCI: the Intramural Research Program of the US National Cancer Institute, NIH, and by support services contracts NO2-CP-11019-50, N02-CP-21013-63 and N02-CP- 65504 with Westat, Inc, Rockville, MD. NICCC: Clalit Health Services in Israel, the Israel Cancer Association and the Breast Cancer Research Foundation (BCRF), NY. NNPIO: the Russian Foundation for Basic Research (grants 17-00-00171, 18-515-45012 and 19-515- 25001). NRG Oncology: U10 CA180868, NRG SDMC grant U10 CA180822, NRG Administrative Office and the NRG Tissue Bank (CA 27469), the NRG Statistical and Data Center (CA 37517) and the Intramural Research Program, NCI. OSU CCG: Ohio State University Comprehensive Cancer Center. PBCS: Italian Association of Cancer Research (AIRC) [IG 2013 N.14477] and Tuscany Institute for Tumors (ITT) grant 2014-2015-2016. SEABASS: Ministry of Science, Technology and Innovation, Ministry of Higher Education (UM.C/HlR/MOHE/06) and Cancer Research Initiatives Foundation. SMC: the Israeli Cancer Association. SWE-BRCA: the Swedish Cancer Society. UCHICAGO: NCI Specialized Program of Research Excellence (SPORE) in Breast Cancer (CA125183), R01 CA142996, 1U01CA161032 and by the Ralph and Marion Falk Medical Research Trust, the Entertainment Industry Fund National Women's Cancer Research Alliance and the Breast Cancer research Foundation. OIO is an ACS Clinical Research Professor. UCLA: Jonsson Comprehensive Cancer Center Foundation; Breast Cancer Research Foundation. UCSF: UCSF Cancer Risk Program and Helen Diller Family Comprehensive Cancer Center. UKFOCR: Cancer Research UK. UPENN: National Institutes of Health (NIH) (R01- CA102776 and R01-CA083855; Breast Cancer Research Foundation; Susan G. Komen Foundation for the cure, Basser Research Center for BRCA. UPITT/MWH: Hackers for Hope Pittsburgh. VFCTG: Victorian Cancer Agency, Cancer Australia, National Breast Cancer Foundation. WCP: Dr Karlan is funded by the American Cancer Society Early Detection Professorship (SIOP-06-258-01-COUN) and the National Center for Advancing Translational Sciences (NCATS), Grant UL1TR000124. This work was supported by the U.S. National Institute of Health (NIH), National Cancer Institute [1RO1CA159868]; as well as by the Breast Cancer Research Foundation; Australia National Health and Medical Research Council [454508, 288704, 145684]; Victorian Health Promotion Foundation; Victorian Breast Cancer Research Consortium; Cancer Australia [1100868, 809195]; National Breast Cancer Foundation [IF 17]; Queensland Cancer Fund; Cancer Councils of New South Wales, Victoria, Tasmania, and South Australia, and Cancer Foundation of Western Australia. We also thank Heather Thorne, Eveline Niedermayr, Sharon Guo, Stephanie Nesci, Lucy Stanhope, Sarah O’Connor, Sandra Picken, the kConFab research nurses and staff, the heads and staff of the Family Cancer Clinics, and the many families who contribute to kConFab. SGBCC was supported by the National Research Foundation Singapore (NRF-NRFF2017-02, awarded to J Li), NUS start-up Grant, National University Cancer Institute Singapore (NCIS) Centre Grant [NMRC/CG/NCIS/2010, NMRC/CG/012/2013, CGAug16M005], Breast Cancer Prevention Programme (BCPP), Asian Breast Cancer Research Fund, and the NMRC Clinician Scientist Award (SI Category) [NMRC/CSA-SI/0015/2017].

KAP is a National Breast Cancer Foundation (Australia) Practitioner Fellow [grant number PRAC-17-004]. M.A. Caligo was supported by Grant 2016 (prog.127/16) from the Fondazione Pisa and by research funding 2017 from the Susan G. Komen Italia onlus.

**Acknowledgements**

All the families and clinicians who contribute to the studies; Catherine M. Phelan for her contribution to CIMBA until she passed away on 22 September 2017; Sue Healey, in particular taking on the task of pathogenic variant classification with the late Olga Sinilnikova; Maggie Angelakos, Judi Maskiell, Gillian Dite, Helen Tsimiklis; members and participants in the New York site of the Breast Cancer Family Registry; members and participants in the Ontario Familial Breast Cancer Registry; Vilius Rudaitis and Laimonas Griškevičius; Drs Janis Eglitis, Anna Krilova and Aivars Stengrevics; Yuan Chun Ding and Linda Steele for their work in participant enrollment and biospecimen and data management; Alicia Barroso, Rosario Alonso and Guillermo Pita; all the individuals and the researchers who took part in CONSIT TEAM (Consorzio Italiano Tumori Ereditari Alla Mammella), in particular: Daniela Zaffaroni, Irene Feroce, Mariarosaria Calvello, Davide Bondavalli, Aliana Guerrieri Gonzaga, Monica Marabelli, A. Viel, Laura Ottini, Giuseppe Giannini, Gabriele Lorenzo Capone, Liliana Varesco, Viviana Gismondi, Maria Grazia Tibiletti, Ileana Carnevali, Antonella Savarese, Aline Martayan, Stefania Tommasi, Brunella Pilato and the personnel of the Cogentech Cancer Genetic Test Laboratory, Milan, Italy. Ms. JoEllen Weaver and Dr. Betsy Bove; The FPGMX group acknowledges members of the Cancer Genetics group (IDIS): Ana Blanco, Marta Santamariña, Miguel Aguado and Belinda Rodríguez-Lage; IFE - Leipzig Research Centre for Civilization Diseases (Markus Loeffler, Joachim Thiery, Matthias Nüchter, Ronny Baber); We thank all participants, clinicians, family doctors, researchers, and technicians for their contributions and commitment to the DKFZ. Genetic Modifiers of Cancer Risk in BRCA1/2 Mutation Carriers (GEMO) study is a study from the National Cancer Genetics Network UNICANCER Genetic Group, France. We wish to pay a tribute to Olga M. Sinilnikova, who with Dominique Stoppa-Lyonnet initiated and coordinated GEMO until she sadly passed away on the 30th June 2014. The team in Lyon (Olga Sinilnikova, Mélanie Léoné, Laure Barjhoux, Carole Verny-Pierre, Sylvie Mazoyer, Francesca Damiola, Valérie Sornin) managed the GEMO samples until the biological resource centre was transferred to Paris in December 2015 (Noura Mebirouk, Fabienne Lesueur, Dominique Stoppa-Lyonnet). We want to thank all the GEMO collaborating groups for their contribution to this study: Coordinating Centre, Service de Génétique, Institut Curie, Paris, France: Muriel Belotti, Ophélie Bertrand, Anne-Marie Birot, Bruno Buecher, Sandrine Caputo, Anaïs Dupré, Emmanuelle Fourme, Marion Gauthier-Villars, Lisa Golmard, Claude Houdayer, Marine Le Mentec, Virginie Moncoutier, Antoine de Pauw, Claire Saule, Dominique Stoppa-Lyonnet, and Inserm U900, Institut Curie, Paris, France: Fabienne Lesueur, Noura Mebirouk.Contributing Centres : Unité Mixte de Génétique Constitutionnelle des Cancers Fréquents, Hospices Civils de Lyon - Centre Léon Bérard, Lyon, France: Nadia Boutry-Kryza, Alain Calender, Sophie Giraud, Mélanie Léone. Institut Gustave Roussy, Villejuif, France: Brigitte Bressac-de-Paillerets, Olivier Caron, Marine Guillaud-Bataille. Centre Jean Perrin, Clermont–Ferrand, France: Yves-Jean Bignon, Nancy Uhrhammer. Centre Léon Bérard, Lyon, France: Valérie Bonadona, Christine Lasset. Centre François Baclesse, Caen, France: Pascaline Berthet, Laurent Castera, Dominique Vaur. Institut Paoli Calmettes, Marseille, France: Violaine Bourdon, Catherine Noguès, Tetsuro Noguchi, Cornel

Popovici, Audrey Remenieras, Hagay Sobol. CHU Arnaud-de-Villeneuve, Montpellier, France: Isabelle Coupier, Pascal Pujol. Centre Oscar Lambret, Lille, France: Claude Adenis, Aurélie Dumont, Françoise Révillion. Centre Paul Strauss, Strasbourg, France: Danièle Muller. Institut Bergonié, Bordeaux, France: Emmanuelle Barouk-Simonet, Françoise Bonnet, Virginie Bubien, Michel Longy, Nicolas Sevenet, Institut Claudius Regaud, Toulouse, France: Laurence Gladieff, Rosine Guimbaud, Viviane Feillel, Christine Toulas. CHU Grenoble, France: Hélène Dreyfus, Christine Dominique Leroux, Magalie Peysselon, Rebischung. CHU Dijon, France: Amandine Baurand, Geoffrey Bertolone, Fanny Coron, Laurence Faivre, Caroline Jacquot, Sarab Lizard. CHU St-Etienne, France: Caroline Kientz, Marine Lebrun, Fabienne Prieur. Hôtel Dieu Centre Hospitalier, Chambéry, France: Sandra Fert Ferrer. Centre Antoine Lacassagne, Nice, France: Véronique Mari. CHU Limoges, France: Laurence Vénat-Bouvet. CHU Nantes, France: Stéphane Bézieau, Capucine Delnatte. CHU Bretonneau, Tours and Centre Hospitalier de Bourges France: Isabelle Mortemousque. Groupe Hospitalier Pitié-Salpétrière, Paris, France: Chrystelle Colas, Florence Coulet, Florent Soubrier, Mathilde Warcoin. CHU Vandoeuvre-les-Nancy, France: Myriam Bronner, Johanna Sokolowska. CHU Besançon, France: Marie-Agnès Collonge- Rame, Alexandre Damette. CHU Poitiers, Centre Hospitalier d’Angoulême and Centre Hospitalier de Niort, France: Paul Gesta. Centre Hospitalier de La Rochelle: Hakima Lallaoui. CHU Nîmes Carémeau, France: Jean Chiesa. CHI Poissy, France: Denise Molina- Gomes. CHU Angers, France : Olivier Ingster; Ilse Coene en Brecht Crombez; Ilse Coene and Brecht Crombez; Alicia Tosar and Paula Diaque; Drs. Sofia Khan, Taru A. Muranen, Carl Blomqvist, Irja Erkkilä and Virpi Palola; The Hereditary Breast and Ovarian Cancer Research Group Netherlands (HEBON) consists of the following Collaborating Centers: Netherlands Cancer Institute (coordinating center), Amsterdam, NL: M.A. Rookus, F.B.L. Hogervorst, F.E. van Leeuwen, M.A. Adank, M.K. Schmidt, D.J. Jenner; Erasmus Medical Center, Rotterdam, NL: J.M. Collée, A.M.W. van den Ouweland, M.J. Hooning, I.A. Boere; Leiden University Medical Center, NL: C.J. van Asperen, P. Devilee, R.B. van der Luijt, T.C.T.E.F. van Cronenburg; Radboud University Nijmegen Medical Center, NL: M.R. Wevers, A.R. Mensenkamp; University Medical Center Utrecht, NL: M.G.E.M. Ausems, M.J. Koudijs; Amsterdam Medical Center, NL: T.A.M. van Os; VU University Medical Center, Amsterdam, NL: K. van Engelen, J.J.P. Gille; Maastricht University Medical Center, NL: E.B. Gómez-Garcia, M.J. Blok, M. de Boer; University of Groningen, NL: L.P.V. Berger, A.H. van der Hout, M.J.E. Mourits, G.H. de Bock; The Netherlands Comprehensive Cancer Organisation (IKNL): S. Siesling, J. Verloop; The nationwide network and registry of histo- and cytopathology in The Netherlands (PALGA): E.C. van den Broek. HEBON thanks the study participants and the registration teams of IKNL and PALGA for part of the data collection; Hong Kong Sanatorium and Hospital; the Hungarian Breast and Ovarian Cancer Study Group members (Janos Papp, Aniko Bozsik, Timea Pocza, Zoltan Matrai, Miklos Kasler, Judit Franko, Maria Balogh, Gabriella Domokos, Judit Ferenczi, Department of Molecular Genetics, National Institute of Oncology, Budapest, Hungary) and the clinicians and patients for their contributions to this study; the Oncogenetics Group (VHIO) and the High Risk and Cancer Prevention Unit of the University Hospital Vall d’Hebron, Miguel Servet Progam (CP10/00617), and the Cellex Foundation for providing research facilities and equipment; the ICO Hereditary Cancer Program team led by Dr. Gabriel Capella; the ICO Hereditary Cancer Program team led by Dr. Gabriel Capella; Catarina Santos and Pedro Pinto; members of the Center of Molecular Diagnosis, Oncogenetics Department and Molecular Oncology Research Center of Barretos Cancer Hospital; Heather Thorne, Eveline Niedermayr, all the kConFab research nurses and staff, the heads and staff of the Family Cancer Clinics, and the Clinical Follow Up Study (which has received funding from the NHMRC, the National Breast Cancer Foundation, Cancer Australia, and the National Institute of Health (USA)) for their contributions to this resource, and the many families who contribute to kConFab; the KOBRA Study Group; Csilla Szabo (National Human Genome Research Institute, National Institutes of Health, Bethesda, MD, USA); Eva Machackova (Department of Cancer Epidemiology and Genetics, Masaryk Memorial Cancer Institute and MF MU, Brno, Czech Republic); and Michal Zikan, Petr Pohlreich and Zdenek Kleibl (Oncogynecologic Center and Department of Biochemistry and Experimental Oncology, First Faculty of Medicine, Charles University, Prague, Czech Republic); Anne Lincoln, Lauren Jacobs; the participants in Hereditary Breast/Ovarian Cancer Study and Breast Imaging Study for their selfless contributions to our research; the NICCC National Familial Cancer Consultation Service team led by Sara Dishon, the lab team led by Dr. Flavio Lejbkowicz, and the research field operations team led by Dr. Mila Pinchev; the investigators of the Australia New Zealand NRG Oncology group; members and participants in the Ontario Cancer Genetics Network; Kevin Sweet, Caroline Craven, Julia Cooper, Amber Aielts, and Michelle O'Conor; Yip Cheng Har, Nur Aishah Mohd Taib, Phuah Sze Yee, Norhashimah Hassan and all the research nurses, research assistants and doctors involved in the MyBrCa Study for assistance in patient recruitment, data collection and sample preparation, Philip Iau, Sng Jen-Hwei and Sharifah Nor Akmal for contributing samples from the Singapore Breast Cancer Study and the HUKM-HKL Study respectively; the Meirav Comprehensive breast cancer center team at the Sheba Medical Center; Christina Selkirk; Håkan Olsson, Helena Jernström, Karin Henriksson, Katja Harbst, Maria Soller, Ulf Kristoffersson; from Gothenburg Sahlgrenska University Hospital: Anna Öfverholm, Margareta Nordling, Per Karlsson, Zakaria Einbeigi; from Stockholm and Karolinska University Hospital: Anna von Wachenfeldt, Annika Lindblom, Brita Arver, Gisela Barbany Bustinza; from Umeå University Hospital: Beatrice Melin, Christina Edwinsdotter Ardnor, Monica Emanuelsson; from Uppsala University: Hans Ehrencrona, Maritta Hellström Pigg, Richard Rosenquist; from Linköping University Hospital: Marie Stenmark-Askmalm, Sigrun Liedgren; Cecilia Zvocec, Qun Niu; Joyce Seldon and Lorna Kwan; Dr. Robert Nussbaum, Beth Crawford, Kate Loranger, Julie Mak, Nicola Stewart, Robin Lee, Amie Blanco and Peggy Conrad and Salina Chan; Simon Gayther, Paul Pharoah, Carole Pye, Patricia Harrington and Eva Wozniak; Geoffrey Lindeman, Marion Harris, Martin Delatycki, Sarah Sawyer, Rebecca Driessen, and Ella Thompson for performing all DNA amplification.SGBCC thanks the participants and all research coordinators for their excellent help with recruitment, data and sample collection.

**Ethics Statement**

**CIMBA Ethics Statement**

All study participants provided written informed consent and participated in research or clinical studies at the host institute under ethically approved protocols. The studies and their approving institutes are: Australian site of the Breast Cancer Family Registry (BCFR-AU) - The University of Melbourne Health Sciences Human Ethics Sub-Committee; Northern California site of the Breast Cancer Family Registry (BCFR-NC) - Northern California Cancer Center Institutional Review Board; New York site of the Breast Cancer Family Registry (BCFR-NY) - Columbia University Medical Center Institutional Review Board; Ontario site of the Breast Cancer Family Registry (BCFR-ON) - Mount Sinai Hospital Research Ethics Board; Philadelphia site of the Breast Cancer Family Registry (BCFR-PA) - Institutional Review Board Fox Chase Cancer Center; Utah site of the Breast Cancer Family Registry (BCFR-UT) - Institutional Review Board University of Utah; Baltic Familial Breast and Ovarian Cancer Consortium (BFBOCC) - Centrālā medicīnas ētikas Komiteja; Lietuvos Bioetikos Komitetas; BRCA-gene mutations and breast cancer in South African women (BMBSA) - University of Pretoria and Pretoria Academic Hospitals Ethics Committee; Beckman Research Institute of the City of Hope (BRICOH) - City of Hope Medical Center Institutional Review Board; Copenhagen Breast Cancer Study (CBCS) - De Videnskabsetiske Komiteer I Region Hovedsladen; Spanish National Cancer Centre (CNIO) - Instituto de Salud Carlos III Comité de Bioética y Bienestar Animal; City of Hope Cancer Center (COH) - City of Hope Institutional Review Board; CONsorzio Studi ITaliani sui Tumori Ereditari Alla Mammella (CONSIT TEAM) - Comitato Etico Indipendente della Fondazione IRCCS "Istituto Nazionale dei Tumori"; National Centre for Scientific Research Demokritos (DEMOKRITOS) - Bioethics committee of NCSR ‘‘Demokritos’’, 240/EHΔ/11.3; National Centre for Scientific Research Demokritos (DEMOKRITOS) - Papageorgiou Hospital Ethics Committee; Dana Farber Cancer Institute (DFCI) - Dana Farber Cancer Institute Institutional Review Board; Deutsches Krebsforschungszentrum (DKFZ) - Ethik-Kommission des

Klinikums der Universität; Deutsches Krebsforschungszentrum (DKFZ) - Hospital Universitario de San Ignacio Comité de Investigaciones y Etica; Deutsches Krebsforschungszentrum (DKFZ) - Shaukat Khanum Memorial Cancer Hospital and Research Centre Institutional Review Board; Epidemiological study of BRCA1 and BRCA2 mutation carriers (EMBRACE) - Anglia & Oxford MREC; Fox Chase Cancer Center (FCCC) - Institutional Review Board Fox Chase Cancer Center; Fundación Pública Galega de Medicina Xenómica - Comite Autonomico de Etica da Investigacion de Galicia; German Consortium of Hereditary Breast and Ovarian Cancer (GC-HBOC) - Ethik-Kommission der Medizinischen Fakultät der Universät zu Köln; Genetic Modifiers of cancer risk in BRCA1/2 mutation carriers (GEMO) - Comité consultatif sur le traitement de I'information en matière de recherche dans le domaine de la santé; Georgetown University (GEORGETOWN) - MedStar Research Institute - Georgetown University Oncology Institutional Review Board; Ghent University Hospital (G-FAST) - Universitair Ziekenhuis Gent - Ethics Committee; Hospital Clinico San Carlos (HCSC) - Comité Ético de Investigación Clínia Hospital Clínico San Carlos; Helsinki Breast Cancer Study (HEBCS) - Helsingin ja uudenmaan sairaanhoitopiiri (Helsinki University Central Hospital ethics committee); HEreditary Breast and Ovarian study Netherlands (HEBON) - Protocol Toetsingscommissie van het Nederlands Kanker Instituut/Antoni van Leeuwenhoek Ziekenhuis; Molecular Genetic Studies of Breast- and Ovarian Cancer in Hungary (HUNBOCS) - Institutional Review Board of the Hungarian National Institute of Oncology; University Hospital Vall d'Hebron (HVH) - The Hospital Universitario Vall d'Hebron Clinical Research Ethics Committee; Institut Català d'Oncologia (ICO) - Catalan Institute of Oncology Institutional Review Board; International Hereditary Cancer Centre (IHCC) - Komisji Bioetycznej Pomorskiej Akademii Medycznej (Pomeranian Medical University Bioethics Committee); Iceland Landspitali - University Hospital (ILUH) - Vísindasiđanefnd National Boethics Committee; Interdisciplinary Health Research International Team Breast Cancer Susceptibility (INHERIT) - Comité d'éthique de la recherche du Centre Hospitalier Universitaire de Québec; Istituto Oncologico Veneto Hereditary Breast and Ovarian Cancer Study (IOVHBOCS) - Centro Oncologico Regionale Azienda Ospedale Di Padova Comitato Etico; Portuguese Oncology Institute-Porto Breast Cancer Study - COMISSÃO DE ÉTICA PARA A SAÚDE (CES) ; Kathleen Cuningham Foundation Consortium for Research into Familial Breast Cancer (KCONFAB) - Queensland Institute of Medical Research - Human Research Ethics Committee; Kathleen Cuningham Foundation Consortium for Research into Familial Breast Cancer (KCONFAB) - Peter MacCallum Cancer Centre Ethics Committee; University of Kansas Medical Center(KUMC) - The University of Kansas Medical Center Human Research Protection Program; Mayo Clinic (MAYO) - Mayo Clinic Institutional Review Boards; McGill University (MCGILL) - McGill Faculty of Medicine Institutional Review Board; Modifier Study of Quantitative Effects on Disease (MOD-SQUAD) - Mayo Clinic Institutional Review Boards; Memorial Sloane Kettering Cancer Center (MSKCC) - Human Biospecimen Utilization Committee; Memorial Sloane Kettering Cancer Center (MSKCC) - Memorial Sloan-Kettering Cancer Center IRB; General Hospital Vienna (MUV) - Ethikkommission der Medizinischen Universität Wien; Women’s College Research Institute Hereditary Breast and Ovarian Cancer Study - University of Toronto Health Sciences Review Ethics Board; National Cancer Institute (NCI) - NIH Ethics Office; National Israeli Cancer Control Center (NICCC) - Carmel Medical Center Institutional Review Board (Helsinki Committee); N.N. Petrov Institute of Oncology (NNPIO) - N.N. Petrov Institional Ethical Committee; NorthShore University HealthSystem (NORTHSHORE) - Institutional Review Board of NorthShore University HealthSystem; NRG Oncology (NRG_ONCOLOGY) - Cancer Prevention and Control Protocol Review Committee; Ontario Cancer Genetics Network (OCGN) - University Health Network Research Ethics Board; The Ohio State University Comprehensive Cancer Center (MACBRCA) - The Ohio State University Cancer Institutional Review Board; Odense University Hospital (OUH) - Den Videnskabsetiske Komité for Region Syddanmark; Pisa Breast Cancer Study (PBCS) - Azienda Ospedaliera Pisana Comitato Etico per lo studio del farmaco sull'uomo; Sheba Medical Centre - Chaim Sheba Medical Center IRB; Swedish Breast Cancer Study (SWE-BRCA) - Regionala Etikprövningsnämnden Stockholm; University of Chicago (UCHICAGO) - The University of Chicago Biological Sciences Division.

Institutional Review Board (BSD IRB); University of California Los Angeles (UCLA) - UCLA Institutional Review Board (UCLA IRB); University of California San Francisco (UCSF) - Human Research Protection Program Institutional Review Board (IRB); UK and Gilda Radner Familial Ovarian Cancer Registries (UKGRFOCR) - Roswell Park Cancer Institute IRB; UK and Gilda Radner Familial Ovarian Cancer Registries (UKGRFOCR) - Cambridge Local Research Ethics Committee; University of Pennsylvania (UPENN) - University of Pennsylvania Institutional Review Board; Cancer Family Registry University of Pittsburg (UPITT) - University of Pittsburgh Institutional Review Board; University of Texas MD Anderson Cancer Center (UTMDACC) - University of Texas MD Anderson Cancer Center Office of Protocol Research Institutional Review Board; Victorian Familial Cancer Trials Group (VFCTG) - Peter MacCallum Cancer Centre Ethics Committee; Women's Cancer Program at Cedars-Sinai Medical Center (WCP) - (Cedars-Sinai Medical Center) CSMC Institutional Review Board.
