## Supplementary figures and images for "Polygenic Risk Modelling for Prediction of Epithelial Ovarian Cancer Risk"

### Supplementary Figure 2

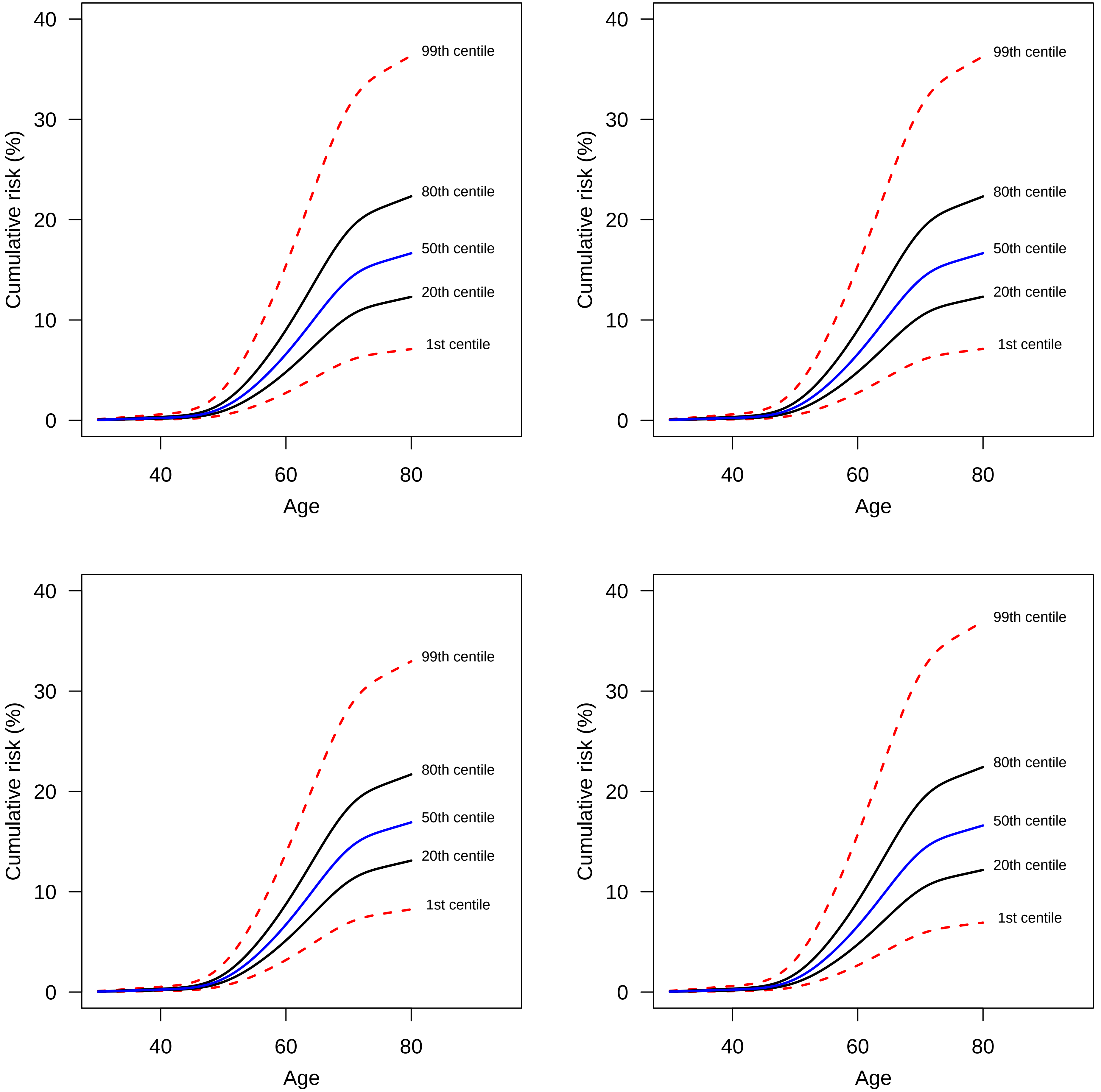

### Supplementary Figure S1

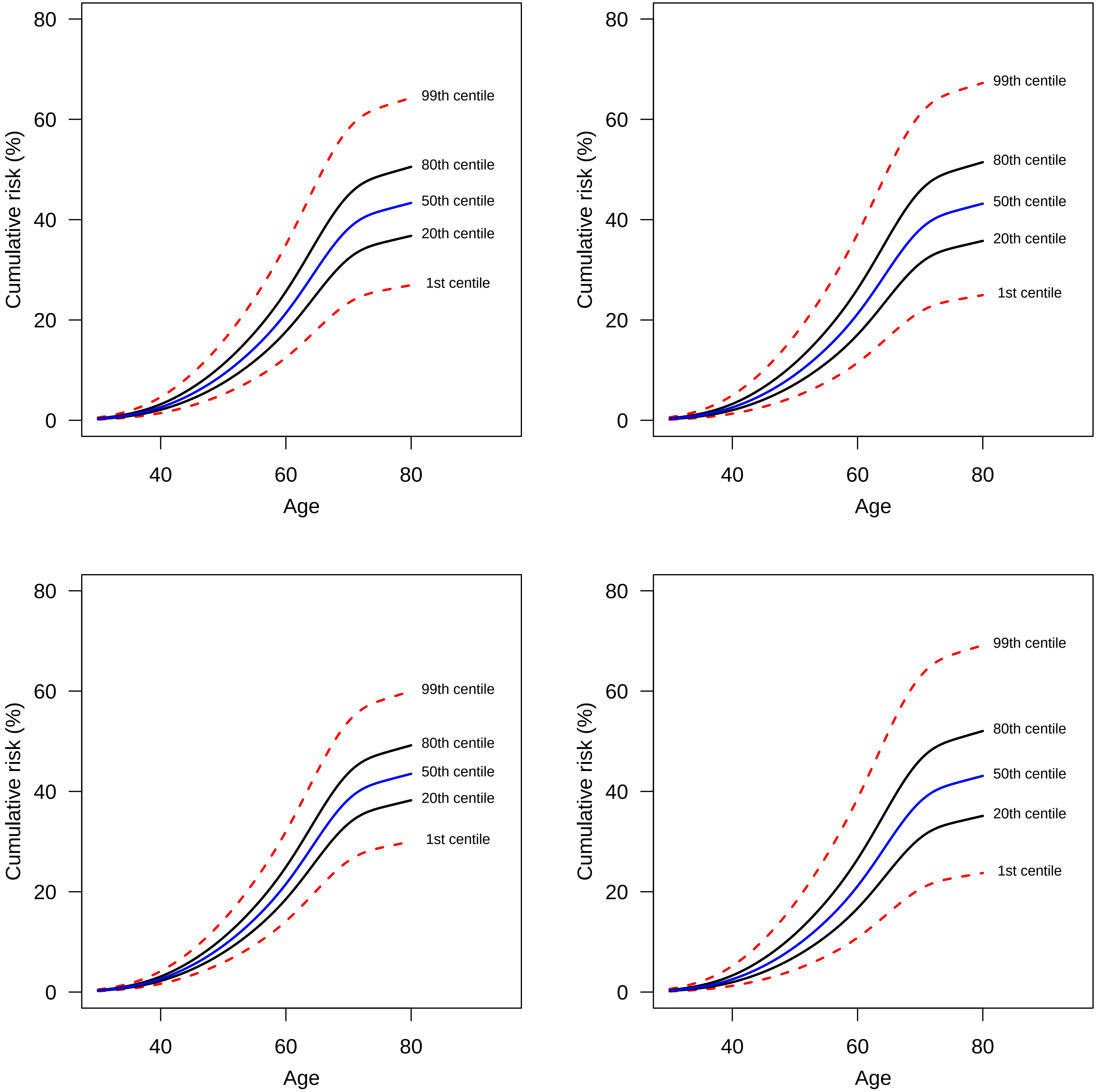
